# Donepezil, galantamine and rivastigmine and progression to memantine add-on therapy in Alzheimer’s disease: a nationwide cohort study emulating a target trial in the Brazilian Unified Health System

**DOI:** 10.64898/2026.11.06.26367201

**Authors:** Gabriela Imbuzeiro Cardoso, Priscilla Vogt Fernandes de Souza, Jhenifer de Paiva Quadros, Mardia Brito de Souza Alcantara, Hércules Rezende Freitas

## Abstract

**Background:** Donepezil, galantamine and rivastigmine are dispensed free of charge to people with mild to moderate Alzheimer’s disease in the Brazilian Unified Health System (SUS), but their comparative long-term effectiveness in routine care is unknown. Under the national clinical protocol, memantine is added to a cholinesterase inhibitor (ChEI) when the disease reaches the moderate stage, so memantine add-on can serve as a marker of clinical progression.

**Methods:** Nationwide new-user cohort study emulating a target trial in which adults with Alzheimer’s disease eligible for a ChEI would be randomly assigned to start donepezil, galantamine or rivastigmine and followed for memantine add-on. We used outpatient medication dispensing authorizations (APAC records of the Outpatient Information System, SIA/SUS) from all 27 federative units, January 2018 to December 2025. We included patients who started a single ChEI between October 2019, after memantine was first recorded nationally, and December 2025, after at least 12 months without anti-dementia dispensing. The outcome was the first memantine dispensing after the index month. Follow-up ended at add-on, recorded death, loss to follow-up (more than 3 observed months without dispensing) or end of data. Deaths were rarely recorded and were treated as censoring. One pseudonymized health card number shared by many patients was excluded, and follow-up ended earlier in the two affected states. We estimated the risk of add-on at 12 to 48 months had all patients started each drug and remained under observation, using stabilized inverse probability of treatment and censoring weights, with 95% confidence intervals (CI) from 200 bootstrap replicates. The analysis assumed no unmeasured confounding or informative censoring given the measured covariates.

**Results:** The cohort included 142 758 new users (donepezil 74 165; galantamine 30 691; rivastigmine 37 902; mean age 77.3 years; 63.0% women), followed for a median of 14 months; 9 812 memantine add-on events occurred. After weighting, all standardized mean differences were below 0.05. The 24-month weighted cumulative incidence of memantine add-on was 9.7% (95% CI 9.4–10.0) with donepezil, 9.0% (8.6–9.4) with galantamine and 5.6% (5.3– 5.9) with rivastigmine. Compared with donepezil, the 24-month risk difference was *−*0.7 percentage points (pp) (*−*1.1 to *−*0.3) for galantamine and *−*4.1 pp (*−*4.5 to *−*3.7) for rivastigmine (risk ratio 0.58, 0.55–0.61). Estimates were stable across sensitivity analyses, including a composite outcome that counted unrecorded deaths and dropouts as events (24month risk difference *−*4.1 pp, *−*4.8 to *−*3.3) and analyses within age groups (risk ratio 0.64 below 75 years). Deaths were almost never recorded in the dispensing data (at most 0.47% of treated patients per year).

**Conclusions:** In routine SUS care, initiation with rivastigmine, mostly as a transdermal patch, was associated with a lower risk of memantine add-on than initiation with donepezil or galantamine. Because disease severity and death are not recorded in the dispensing data, this difference may reflect prescribing patterns or residual confounding by severity rather than slower progression; additional analyses made differential mortality a less likely explanation. It should be confirmed with linked clinical and mortality data.

## 1 Introduction

### Background

The number of people living with dementia worldwide was estimated at 57.4 million in 2019 and is projected to reach 152.8 million in 2050, with the largest relative increases in North Africa and the Middle East and in eastern sub-Saharan Africa [1]. In Brazil, the national dementia report estimated a prevalence of 8.5% among people aged 60 years or older in 2019 [2]. Alzheimer’s disease is the most common cause.

Cholinesterase inhibitors (ChEIs) are the standard symptomatic treatment for mild to moderate Alzheimer’s disease, and the 2024 Lancet Commission recommends that ChEIs and memantine be provided to people with the disease [3]. Placebo-controlled trials, most lasting about 6 months, show modest benefits on cognition, global function and activities of daily living for galantamine [4], donepezil [5] and rivastigmine, including the transdermal patch [6]. The same Cochrane review found no evidence of differences in efficacy between the three drugs [4]; the few head-to-head trials gave conflicting results, and adjusted indirect comparisons found no significant differences in cognition [7]. Evidence on long-term outcomes in routine care comes mainly from registries in high-income countries [8].

In the Brazilian Unified Health System (SUS), the three ChEIs are dispensed free of charge through the Specialized Component of Pharmaceutical Assistance, under a national clinical protocol that defines eligibility by cognitive and functional staging [9]; they have been provided since the first version of the protocol, in 2002 [10]. Memantine was incorporated in 2017 for moderate disease, in combination with a ChEI, and for severe disease as monotherapy [9, 11]. Each dispensing is authorized and recorded monthly in a national information system, which makes it possible to follow every treated patient over time. Under the protocol, the addition of memantine to a ChEI corresponds to clinical progression to the moderate stage, so time to memantine add-on is a pragmatic marker of progression that is recorded for all patients. An earlier Brazilian study described failures in monitoring compliance with the protocol at dispensing [12], but no study has compared the three ChEIs nationwide. A randomized trial comparing the three drugs over several years in routine care is unlikely to be conducted, because all three are approved, off patent and available in the SUS. Emulating such a trial with the national dispensing records makes explicit the comparison of interest and helps avoid common design errors, such as immortal time and the inclusion of prevalent users [13, 14]. The specification of the target trial (eligibility, treatment strategies and outcome) was informed by the placebo-controlled trials of the three ChEIs [4–6] and by the national clinical protocol [9].

### Objectives

The primary objective was to estimate the effect of starting donepezil, galantamine or rivastigmine on the risk of memantine add-on up to 48 months, in patients with Alzheimer’s disease treated in the SUS. The causal question was: among patients eligible for a ChEI under the national protocol, how would the cumulative risk of memantine add-on differ if all had started galantamine or rivastigmine instead of donepezil, regardless of later switching or discontinuation (intention-to-treat effect)? The prespecified hypothesis was that the three drugs would not differ, in line with trial evidence. A secondary objective was to assess whether nationwide SUS dispensing records are fit for this purpose, by examining identifier stability, completeness and the recording of deaths before the analysis.

## 2 Methods

### 2.1 Study design

This is a retrospective cohort study of new users of ChEIs, designed to emulate a hypothetical pragmatic randomized trial (target trial) [13, 14] with an active-comparator, new-user design [15, 16]. Table 1 summarizes the protocol of the target trial and its emulation. The study is reported following the STROBE statement [17, 18] (checklist in Supplementary Table S1) and the TARGET guideline [19] (checklist in Supplementary Table S2), and takes into account the RECORD and RECORD-PE extensions for routinely collected data [20, 21]: the codes used to identify drugs and diagnoses are given below, no record linkage with other databases was performed, and data cleaning is described in the data quality section.

**Table 1:**
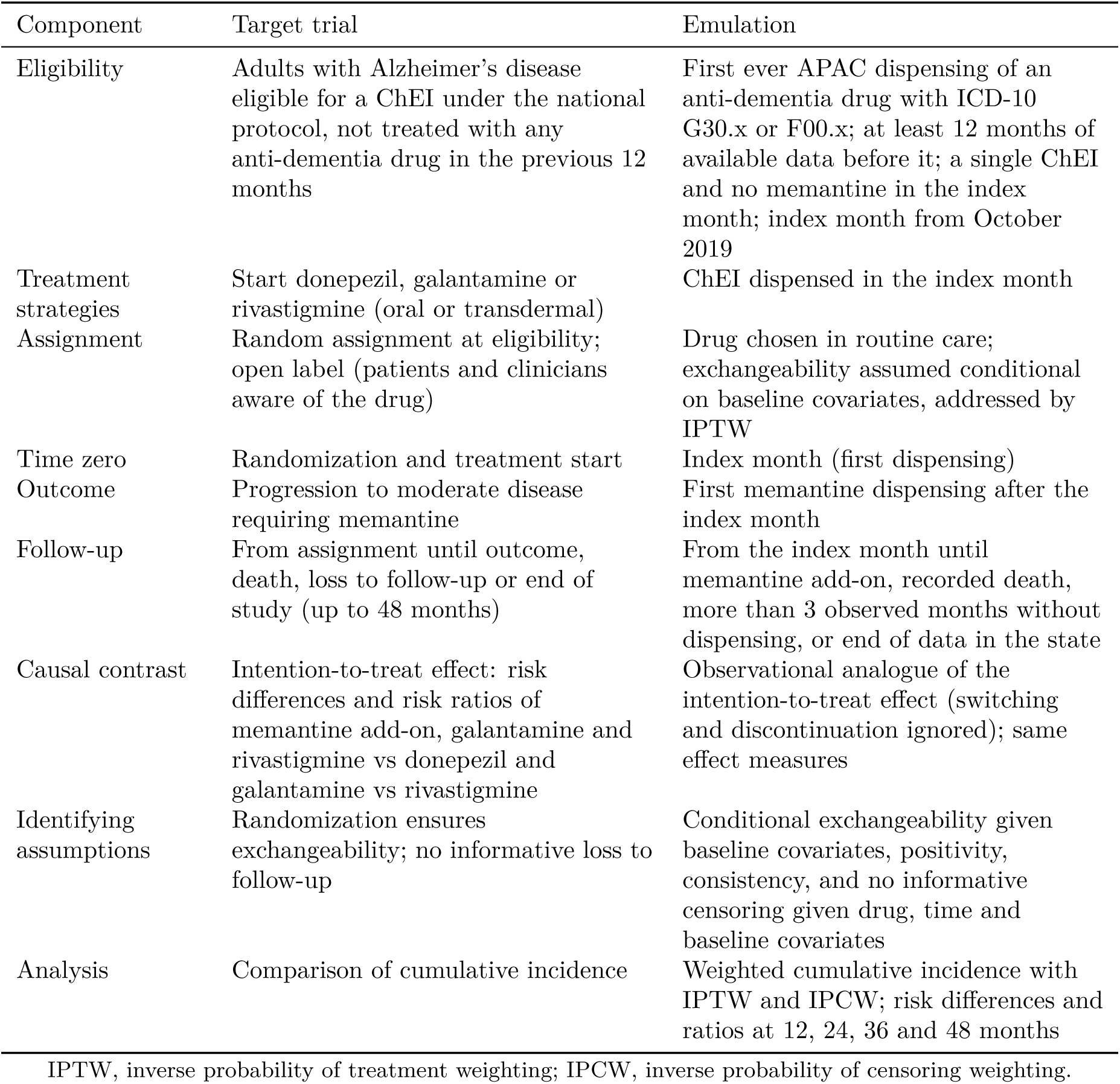
Specification of the target trial and its emulation with SIA/SUS dispensing records.

### 2.2 Setting

The SUS provides universal, free health care to the Brazilian population of about 203 million people. Medicines for Alzheimer’s disease are dispensed by state pharmacies of the Specialized Component of Pharmaceutical Assistance after a specialist prescription and an authorization that is renewed every three months; each monthly dispensing generates a record in the Outpatient Information System (SIA/SUS). Data covered dispensings from January 2018 to December 2025 in all 27 federative units. Recruitment into the cohort ran from October 2019 to December 2025, and follow-up ended in December 2025 (October 2024 in Goiás and July 2025 in São Paulo; see below). Data were downloaded between September 28 and 30, 2026.

### 2.3 Participants

Monthly records were aggregated into person-months. The index month was the first month with any dispensing of donepezil, galantamine, rivastigmine or memantine. Eligible patients had a main diagnosis of Alzheimer’s disease (ICD-10 G30.x or F00.x) and an index month at least 12 months after the start of data (January 2018), which ensured at least 12 months without anti-dementia dispensing. Because memantine was first recorded nationally in September 2019, cohort entry started in October 2019, so that the outcome and the exclusion of memantine users at baseline were observable for every participant. Patients with memantine or more than one ChEI in the index month were excluded. The APAC type (initial or continuation) was not used to define new users because initial authorizations recur at every quarterly renewal (Supplementary Table S3). Follow-up methods are described under Outcome and follow-up.

### 2.4 Variables

#### Exposure

The ChEI dispensed in the index month: donepezil (5 or 10 mg tablets), galantamine (8, 16 or 24 mg extended-release capsules) or rivastigmine (capsules, oral solution or 9 or 18 mg transdermal patch), identified by SIGTAP procedure codes.

#### Outcome

Memantine add-on, defined as the first memantine dispensing in any month after the index month.

#### Potential confounders

Age, sex, race/colour, ICD-10 code of the main diagnosis, body mass index (BMI), state of residence and calendar month of initiation, all measured in the index month. No effect modifiers were prespecified.

### 2.5 Data sources and measurement

We used the medication records of SIA/SUS (file family “AM”), published monthly by the Department of Informatics of SUS (DATASUS) for each federative unit. Each record corresponds to a high-complexity outpatient procedure authorization (APAC) for a dispensed medicine and contains the processing and competence months, the SIGTAP code of the medicine, the ICD-10 code of the main diagnosis, age, sex, race/colour (self-reported or reported by staff), municipality of residence, the dispensing establishment, weight and height, the exit reason and a death indicator. The patient’s national health card number is present in an obfuscated form that is stable across records.

All AM files were downloaded with the development version of the microdatasus package [22], keeping records with ICD-10 G30.x or F00.x. The health card number was replaced immediately by a pseudonymous identifier computed as a keyed hash (HMAC-SHA256) with a secret key stored outside the project, and the original value was discarded. Procedure codes were mapped to drugs with the SIGTAP reference table. Age was taken from the record (patients aged 100 years or more were coded as 100); BMI was calculated from recorded weight and height when both were within plausible ranges (25–250 kg and 100–220 cm). The same definitions were applied to the three groups.

### 2.6 Bias

Selection and immortal time bias were avoided by aligning eligibility, treatment assignment and the start of follow-up at the index month, and by restricting to new users with an active comparator [13, 16]. Confounding by measured baseline characteristics was addressed by IPTW. Informative loss to follow-up, including unrecorded deaths, was addressed by IPCW. Two data problems could bias follow-up and were handled explicitly after a pilot study and national quality checks: unavailable monthly files and a health card number shared by many patients (Section 2.8). Residual confounding by disease severity, which is not recorded, is discussed in the limitations.

### 2.7 Study size

All eligible patients in the national data were included; no sample size was calculated in advance. A pilot in four states (Rio de Janeiro, Minas Gerais, Bahia and Rio Grande do Sul) had shown that each group would include more than 200 new users and more than 30 events, the minimum we set for the national analysis; the precision of the national estimates is reflected in the confidence intervals.

### 2.8 Data quality assessment and handling

Before the analysis we checked file availability, record counts by state and month, processing delays, missing values, duplicate records, monthly and year-to-year retention of identifiers, coding of procedures and diagnoses, recording of deaths, completeness of covariates and positivity, first in the pilot states and then nationally. The criteria and their assessment are given in Supplementary Table S5, and the full results of the checks in Supplementary Tables S6 to S19. The full pipeline, including the checks, is available as reproducible code (see Data availability).

Two problems required changes to the design. First, 29 of the 2 592 state-month files were not available on DATASUS; these months were treated as unobserved, so a gap in dispensing that overlapped an unavailable file did not count towards loss to follow-up. Second, one pseudonymous identifier had up to 2 444 records per month, which is impossible for a single patient and indicates a health card number shared by many patients. Any identifier with more than 20 records in one month was classified as a placeholder and its records were excluded. For each state, the end of data was set to the month before the share of placeholder records first exceeded 5%; in a sensitivity analysis the threshold was 1%.

### 2.9 Outcome and follow-up

Follow-up started in the index month and ended at memantine add-on, recorded death (exit reason 41–43 or death indicator), loss to follow-up (more than 3 observed months without any dispensing before the end of data) or the end of data in the state of the last dispensing. Because deaths were very rarely recorded (see Results), death was not treated as a competing event; recorded deaths and losses were handled as censoring that may be informative.

### 2.10 Quantitative variables

Age and calendar month of initiation were modelled as continuous variables with natural cubic splines (3 and 4 degrees of freedom). BMI was grouped as *<*18.5, 18.5–24.9, 25.0–29.9 and *≥*30 kg/m^2^, plus a missing category, to accommodate missing values and non-linearity. For description, age was grouped as *<*65, 65–74, 75–84 and *≥*85 years. Race/colour categories with fewer than 30 patients and states with fewer than 50 new users were pooled; Amapá, with too few new users per drug, was pooled with the neighbouring state of Pará.

### 2.11 Statistical methods

#### Estimands and identifying assumptions

For each of the three pairwise contrasts, the estimands were the differences and ratios of the cumulative risk of memantine add-on at 12, 24, 36 and 48 months had all eligible patients started one drug rather than the other, in the hypothetical absence of censoring (observational analogue of the intention-to-treat effect). Identification relied on four assumptions: conditional exchangeability, that is, no unmeasured confounding of the drug choice given the measured baseline covariates (age, sex, race/colour, ICD-10 code, BMI, state of residence and calendar month); positivity, that is, a non-zero probability of starting each drug in every covariate stratum, checked by the distribution of propensity scores and weights; consistency, that is, a well-defined intervention, supported by the restriction to patients starting a single ChEI in the index month; and no informative censoring given drug, time since initiation and the same baseline covariates. All covariates were measured in the index month from the APAC record, as described under Data sources and measurement. Correct specification of the treatment and censoring models was also assumed.

#### Confounding

Treatment weights were estimated with a multinomial logistic model for the three drugs [23, 24], including all baseline covariates listed above. Stabilized weights were the marginal probability of the drug received divided by its predicted probability [25, 26]. Covariate balance was assessed with standardized mean differences for all pairwise comparisons, with 0.1 as the threshold for meaningful imbalance [27, 28].

#### Outcome analysis

Censoring by loss to follow-up or recorded death was modelled with pooled logistic regression on person-months, including drug, time since initiation (natural spline, 4 degrees of freedom) and baseline covariates in the denominator and drug and time in the numerator [29]. Administrative end of data was treated as non-informative. The product of treatment and censoring weights was truncated at the 1st and 99th percentiles. Cumulative incidence of memantine add-on was estimated as one minus the weighted Kaplan–Meier estimator on person-month data [30]. We report cumulative incidence, risk differences and risk ratios at 12, 24, 36 and 48 months, rather than hazard ratios [31]. Percentile 95% CIs were obtained from 200 non-parametric bootstrap replicates that resampled patients and refitted all models [32]. Crude (unweighted) estimates were obtained with the Kaplan–Meier estimator, with Greenwood standard errors and delta-method CIs for differences and ratios.

#### Subgroups and interactions

Analyses within age groups and by rivastigmine formulation were done as additional analyses (see below); no formal tests of interaction were performed.

#### Missing data

Age, sex, ICD-10 code and state of residence were complete. Race/colour was not informed for 22.9% and BMI was missing for 2.7% of patients; both were analysed with an explicit missing category. No imputation was done.

#### Loss to follow-up

Patients without dispensing for more than 3 observed months were censored at their last dispensing, and IPCW was used to account for the dependence of this censoring on drug, time and baseline covariates.

#### Sensitivity analyses

We repeated the main analysis with untruncated weights, without censoring weights, without any weighting, and in the cohort defined with the stricter 1% placeholder threshold.

#### Additional analyses on disease severity and unrecorded deaths

Because severity and death are not recorded, three additional analyses examined alternative explanations for differences between drugs. First, to address differential mortality and dropout, we used a composite outcome of memantine add-on or end of dispensing (loss to follow-up or recorded death, dated at the first month without dispensing), whichever came first; unrecorded deaths and dropouts are events in this analysis, so they cannot favour one drug. It was weighted by stabilized IPTW only, because the only remaining censoring was the administrative end of data, with 95% CIs from 200 bootstrap replicates. Second, to address the selection of frailer patients for the transdermal patch, oral and transdermal rivastigmine users were analysed separately, each together with all donepezil and galantamine users. Third, because unrecorded deaths are much more frequent at older ages, the main analysis was repeated within age groups (*<*75, 75–84 and *≥*85 years). In the second and third analyses the propensity and censoring models were refitted in each subset. Finally, we calculated E-values, the minimum strength of association on the risk ratio scale that an unmeasured confounder, such as disease severity, would need with both the drug and the outcome to explain away an estimate [33].

Analyses were done in R 4.5.3 [34] with the packages data.table, survival [35] and nnet [36], in a single Quarto document that reproduces the whole pipeline.

### 2.12 Ethics

The study used publicly available, de-identified secondary data. Under Resolution 510/2016 of the Brazilian National Health Council, research that uses information of public access or in the public domain is not registered or evaluated by the CEP/CONEP system (article 1, sole paragraph, items II and III) [37]; ethics committee approval was therefore not required.

## 3 Results

### 3.1 Participants

The national files contained 7 785 263 dispensing records with an Alzheimer’s diagnosis, corresponding to 291 910 pseudonymous identifiers; 2 563 of the 2 592 expected state-month files were available. All procedure codes mapped to one of the four drugs. The placeholder identifier carried 160 183 records (2.1%), almost all from Goiás and São Paulo (Supplementary Figure S1A). In Goiás it carried 99.4–99.6% of records from November 2024 onwards; in São Paulo its share rose from 0.7% in November 2024 to 3.8% in July 2025 and 45–95% from September 2025. The end of data was therefore set to October 2024 in Goiás and July 2025 in São Paulo (November 2024 with the 1% threshold).

Figure 1 shows the number of patients at each stage of selection and the reasons for exclusion. Of 290 332 patients with at least one eligible record, 142 758 were new users of a single ChEI from October 2019: 74 165 (52.0%) started donepezil, 30 691 (21.5%) galantamine and 37 902 (26.6%) rivastigmine, of whom 29 792 (78.6%) started with the transdermal patch.

**Figure 1:**
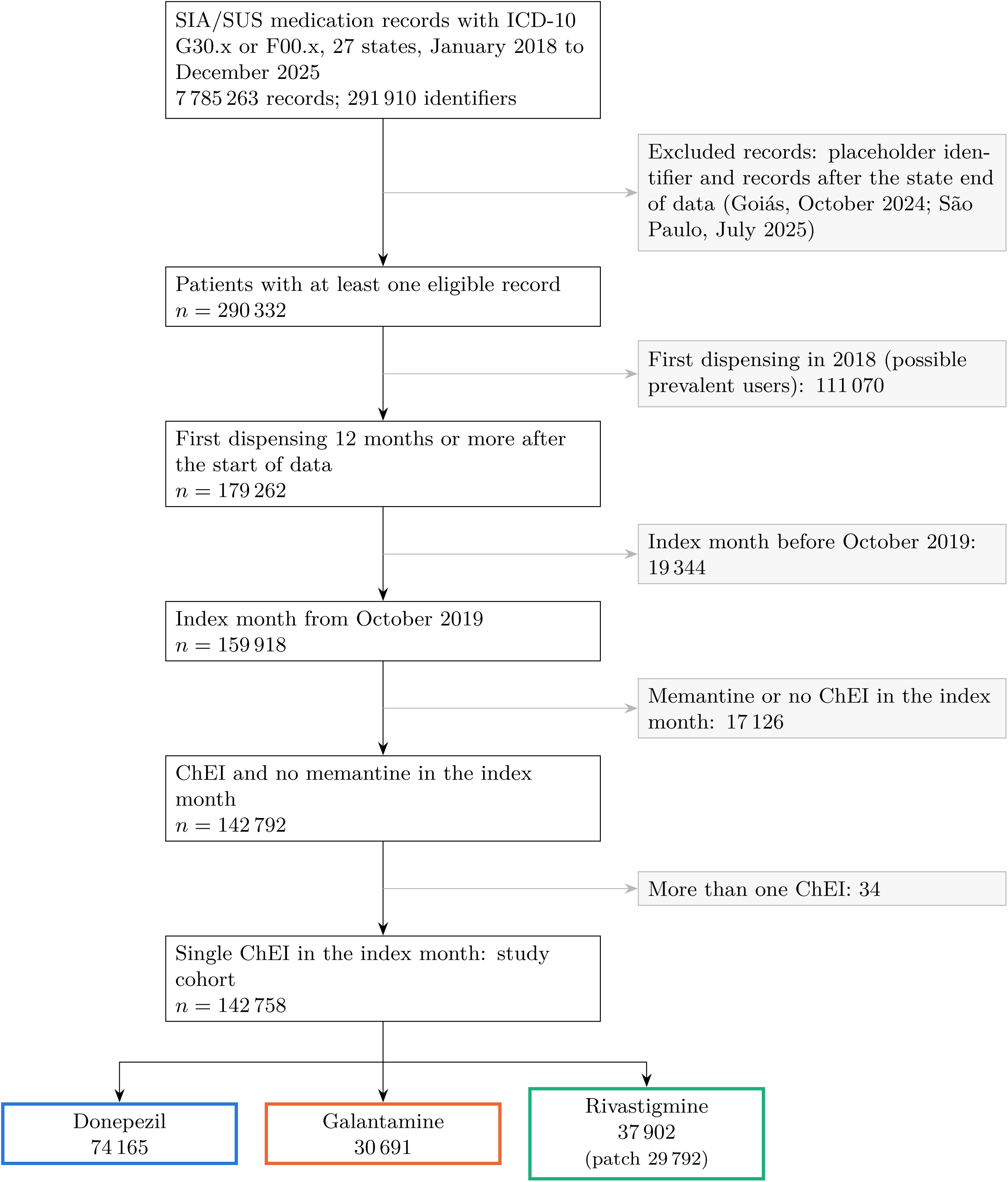
Selection of the study cohort, with the number of patients at each stage and the reasons for exclusion.

### 3.2 Descriptive data

Table 2 shows baseline characteristics, including the number of patients with missing data for each variable. Rivastigmine users were older (mean 78.3 vs 77.0 and 76.7 years) and more often women (66.9% vs 61.8% and 61.1%). The largest differences between drugs were in state of residence and calendar time of initiation. São Paulo (48.3% of the cohort) and Paraná (15.1%) contributed most new users. Participants contributed 2 773 286 person-months (231 107 personyears), with a median follow-up of 14 months (interquartile range 5–29) in each group. Follow-up ended by loss in 43.5% of patients, by administrative end of data in 48.9% and by recorded death in 0.8%.

**Table 2:** Baseline characteristics and follow-up of new users of cholinesterase inhibitors, SUS, October 2019 to December 2025.

|  | Donepezil | Galantamine | Rivastigmine | Total |
|---|---|---|---|---|
| New users, *n* | 74 165 | 30 691 | 37 902 | 142 758 |
| Age, mean (SD), years | 77.0 (8.6) | 76.7 (8.3) | 78.3 (8.3) | 77.3 (8.5) |
| <65 | 8.2 | 7.9 | 5.8 | 7.5 |
| 65–74 | 27.1 | 28.3 | 23.5 | 26.4 |
| 75–84 | 45.8 | 46.5 | 47.3 | 46.3 |
| ≥85 | 18.9 | 17.3 | 23.3 | 19.7 |
| Women | 61.8 | 61.1 | 66.9 | 63.0 |
| Race/colour |  |  |  |  |
| White | 53.0 | 53.4 | 52.1 | 52.8 |
| Brown (*parda*) | 19.4 | 20.7 | 19.4 | 19.7 |
| Black | 3.5 | 3.0 | 2.0 | 3.0 |
| Asian or Indigenous | 1.5 | 1.8 | 1.9 | 1.7 |
| Not informed (missing) | 22.7 | 21.1 | 24.7 | 22.9 |
| ICD-10 code |  |  |  |  |
| G30.0 | 53.4 | 56.2 | 52.7 | 53.8 |
| G30.1 | 24.2 | 23.7 | 29.2 | 25.4 |
| G30.8 | 9.3 | 8.7 | 8.2 | 8.9 |
| F00.0–F00.2 | 13.1 | 11.4 | 9.9 | 11.9 |
| Body mass index, mean (SD), kg/m<sup>2</sup> | 25.6 (4.5) | 25.8 (4.5) | 25.1 (4.4) | 25.5 (4.5) |
| Missing | 2.4 | 2.9 | 3.0 | 2.7 |
| Region of residence |  |  |  |  |
| North | 1.3 | 1.4 | 1.8 | 1.5 |
| Northeast | 10.0 | 13.2 | 14.3 | 11.8 |
| Southeast | 61.3 | 57.7 | 57.7 | 59.6 |
| South | 24.6 | 23.5 | 21.0 | 23.4 |
| Center-West | 2.8 | 4.2 | 5.2 | 3.7 |
| Year of initiation |  |  |  |  |
| 2019 (October–December) | 4.5 | 3.5 | 5.2 | 4.5 |
| 2020–2021 | 28.2 | 25.5 | 31.6 | 28.5 |
| 2022–2023 | 35.9 | 37.6 | 29.9 | 34.7 |
| 2024–2025 | 31.5 | 33.4 | 33.3 | 32.4 |
| Transdermal formulation | – | – | 78.6 | – |
| Person-years of follow-up | 118 910 | 49 074 | 63 124 | 231 107 |
| Follow-up, median (IQR), months | 14 (5–29) | 14 (5–29) | 14 (5–30) | 14 (5–29) |
| End of follow-up |  |  |  |  |
| Memantine add-on | 7.4 | 7.8 | 5.1 | 6.9 |
| Loss to follow-up | 44.8 | 40.8 | 43.1 | 43.5 |
| Recorded death | 0.8 | 0.7 | 0.7 | 0.8 |
| End of data | 46.9 | 50.7 | 51.1 | 48.9 |

Values are percentages unless stated otherwise. SD, standard deviation; IQR, interquartile range. Age, sex, ICD-10 code and state of residence had no missing values. ICD-10 codes as recorded in the APAC. State of residence (26 units, Amapá pooled with Pará) was included in the models; regions are shown for brevity.

### 3.3 Outcome data

There were 9 812 memantine add-on events: 5 479 with donepezil, 2 391 with galantamine and 1 942 with rivastigmine, corresponding to crude rates of 46.1, 48.7 and 30.8 per 1 000 personyears. The numbers of patients at risk over time are shown in Figure 2A, and crude cumulative incidence in Table 3.

**Figure 2:**
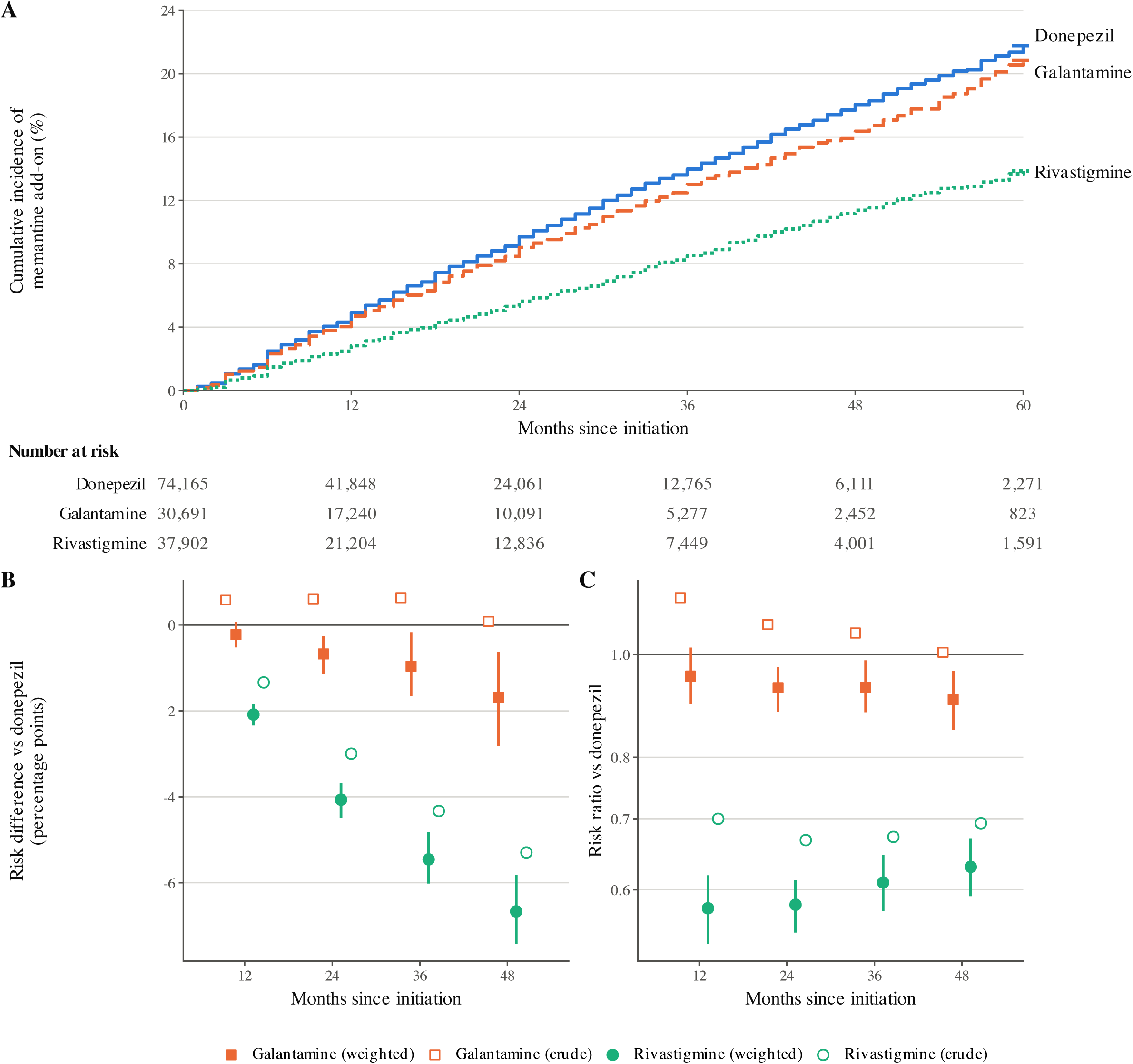
Memantine add-on after initiation of donepezil, galantamine or rivastigmine. (A) Weighted cumulative incidence (inverse probability of treatment and censoring weights) and number of patients at risk. (B) Risk difference and (C) risk ratio relative to donepezil, weighted (filled symbols, 95% bootstrap CI) and crude (open symbols); the risk ratio is shown on a logarithmic scale.

**Table 3:** Crude and weighted cumulative incidence of memantine add-on and comparisons between drugs.

|  |  | 12 months | 24 months | 36 months | 48 months |
|---|---|---|---|---|---|
| *Cumulative incidence, % (95% CI)* |  |  |  |  |  |
| Donepezil | Crude | 4.5 (4.3, 4.6) | 9.0 (8.7, 9.3) | 13.2 (12.8, 13.6) | 17.3 (16.7, 17.8) |
|  | Weighted | 4.9 (4.8, 5.1) | 9.7 (9.4, 10.0) | 14.0 (13.6, 14.4) | 18.0 (17.5, 18.6) |
| Galantamine | Crude | 5.0 (4.8, 5.3) | 9.6 (9.2, 10.1) | 13.9 (13.3, 14.5) | 17.3 (16.5, 18.2) |
|  | Weighted | 4.7 (4.5, 5.0) | 9.0 (8.6, 9.4) | 13.0 (12.5, 13.6) | 16.4 (15.6, 17.3) |
| Rivastigmine | Crude | 3.1 (2.9, 3.3) | 6.0 (5.7, 6.4) | 8.9 (8.5, 9.4) | 12.0 (11.3, 12.6) |
|  | Weighted | 2.8 (2.6, 3.0) | 5.6 (5.3, 5.9) | 8.5 (8.1, 9.0) | 11.4 (10.8, 12.0) |
| *Risk difference, per 100 patients (95% CI)* |  |  |  |  |  |
| GAL vs DON | Crude | 0.6 (0.2, 0.9) | 0.6 (0.1, 1.1) | 0.6 (−0.1, 1.4) | 0.1 (−0.9, 1.1) |
|  | Weighted | −0.2 (−0.5, 0.1) | −0.7 (−1.1, −0.3) | −1.0 (−1.7, −0.2) | −1.7 (−2.8, −0.6) |
| RIV vs DON | Crude | −1.3 (−1.6, −1.1) | −3.0 (−3.4, −2.6) | −4.3 (−4.9, −3.7) | −5.3 (−6.1, −4.5) |
|  | Weighted | −2.1 (−2.3, −1.8) | −4.1 (−4.5, −3.7) | −5.5 (−6.0, −4.8) | −6.7 (−7.4, −5.8) |
| GAL vs RIV | Crude | 1.9 (1.6, 2.3) | 3.6 (3.1, 4.2) | 5.0 (4.2, 5.7) | 5.4 (4.4, 6.4) |
|  | Weighted | 1.9 (1.5, 2.2) | 3.4 (2.8, 3.9) | 4.5 (3.7, 5.3) | 5.0 (4.0, 6.0) |
| *Risk ratio (95% CI)* |  |  |  |  |  |
| GAL vs DON | Crude | 1.13 (1.06, 1.21) | 1.07 (1.01, 1.13) | 1.05 (0.99, 1.10) | 1.00 (0.95, 1.06) |
|  | Weighted | 0.95 (0.90, 1.01) | 0.93 (0.88, 0.97) | 0.93 (0.88, 0.99) | 0.91 (0.85, 0.96) |
| RIV vs DON | Crude | 0.70 (0.65, 0.76) | 0.67 (0.63, 0.71) | 0.67 (0.63, 0.71) | 0.69 (0.65, 0.74) |
|  | Weighted | 0.58 (0.53, 0.62) | 0.58 (0.55, 0.61) | 0.61 (0.57, 0.65) | 0.63 (0.59, 0.67) |
| GAL vs RIV | Crude | 1.62 (1.48, 1.76) | 1.60 (1.49, 1.71) | 1.56 (1.46, 1.67) | 1.45 (1.35, 1.55) |
|  | Weighted | 1.66 (1.51, 1.82) | 1.60 (1.48, 1.72) | 1.53 (1.42, 1.64) | 1.44 (1.32, 1.55) |

DON, donepezil; GAL, galantamine; RIV, rivastigmine. Crude: Kaplan–Meier with Greenwood standard errors and delta-method CIs. Weighted: stabilized inverse probability of treatment and censoring weights, truncated at the 1st and 99th percentiles, adjusted for age, sex, race/colour, ICD-10 code, body mass index, state of residence and calendar month; 95% CI from 200 bootstrap replicates (percentile method).

### 3.4 Main results

Before weighting, the largest absolute standardized mean differences were for state of residence (up to 0.23), age (0.20), ICD-10 G30.1, sex and calendar time. Stabilized treatment weights had a mean close to 1 in each group (1st to 99th percentile 0.44 to 2.07; maximum 6.12); after truncation of the combined weights, the effective sample sizes were 71 230, 29 122 and 34 294. After weighting, all standardized mean differences were below 0.05 (maximum 0.041; Figure 3A; Supplementary Table S19).

**Figure 3:**
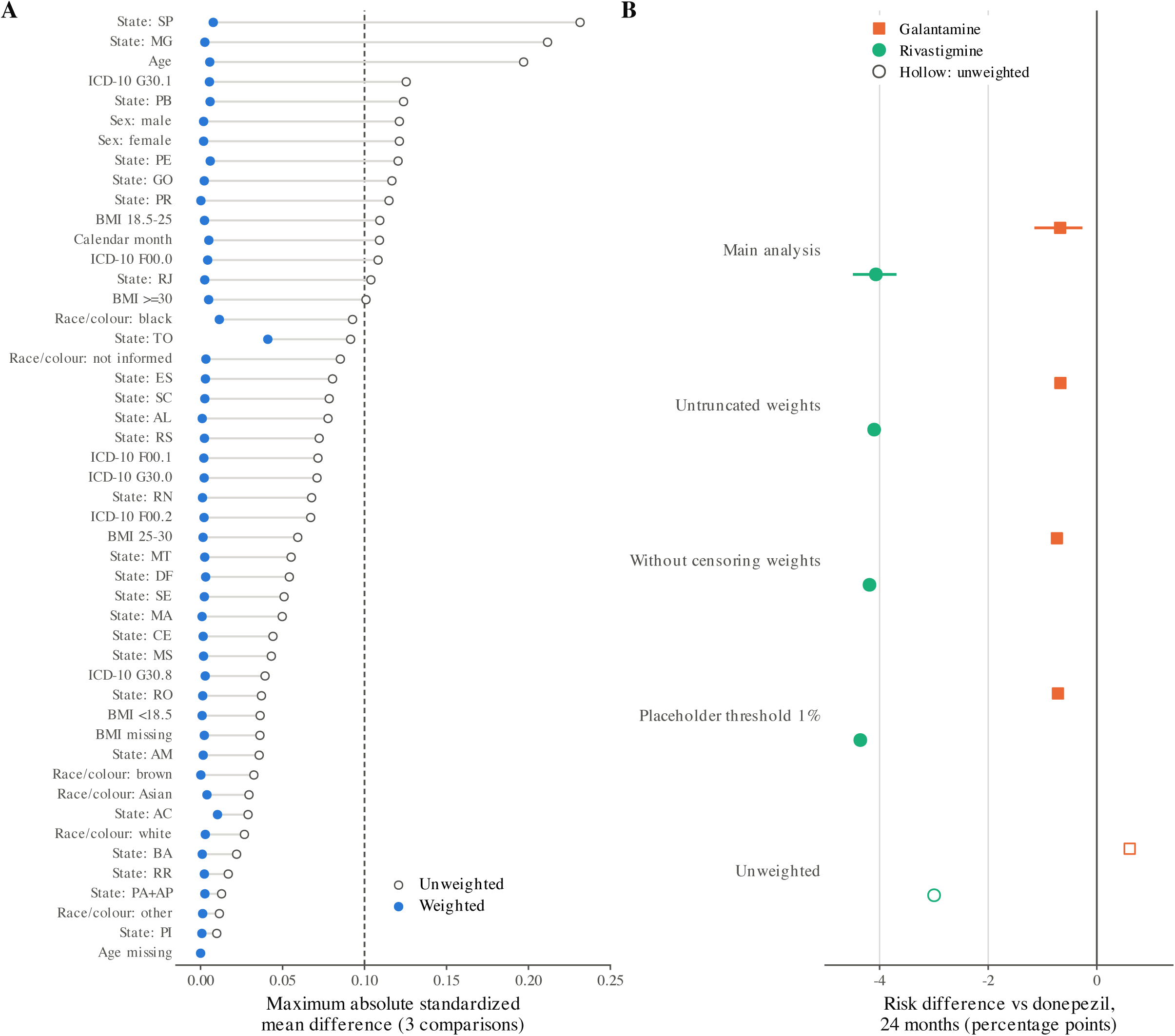
(A) Covariate balance before (open circles) and after (filled circles) weighting: maximum absolute standardized mean difference over the three pairwise comparisons; dashed line, 0.1. (B) Risk difference relative to donepezil at 24 months in the main and sensitivity analyses; the horizontal line in the main analysis is the 95% bootstrap CI; open symbols, unweighted.

Table 3 and Figure 2 show crude and weighted estimates. The weighted estimates were adjusted for age, sex, race/colour, ICD-10 code, BMI category, state of residence and calendar month, and for informative censoring. At 24 months the weighted cumulative incidence of memantine add-on was 9.7% with donepezil, 9.0% with galantamine and 5.6% with rivastigmine; at 48 months it was 18.0%, 16.4% and 11.4%. In absolute terms, compared with donepezil, rivastigmine was associated with 4.1 fewer add-ons per 100 patients at 24 months (risk ratio 0.58, 95% CI 0.55–0.61) and 6.7 fewer per 100 at 48 months (0.63, 0.59–0.67). Galantamine was associated with a small reduction relative to donepezil after weighting (*−*0.7 per 100 at 24 months, *−*1.1 to *−*0.3; *−*1.7 per 100 at 48 months, *−*2.8 to *−*0.6), whereas the crude estimates showed a small increase (Table 3).

### 3.5 Other analyses

Estimates were similar with untruncated weights, without censoring weights, and in the cohort defined with the 1% placeholder threshold (134 531 patients) (Figure 3B; Supplementary Table S4). The rivastigmine versus donepezil difference at 24 months ranged from *−*4.1 to *−*4.4 per 100 across weighted analyses and was *−*3.0 without weighting. The galantamine versus donepezil difference changed sign without weighting (+0.6 per 100), which shows that it depends on the adjustment for baseline differences.

The lower risk with rivastigmine persisted in the additional analyses (Supplementary Tables S20 and S21). With the composite outcome of memantine add-on or end of dispensing, which counts unrecorded deaths and dropouts as events, rivastigmine was associated with 4.1 fewer events per 100 patients than donepezil at 24 months (95% CI 3.3 to 4.8; risk ratio 0.92, 0.90–0.93), and galantamine with 2.2 fewer (1.4 to 2.9). The relative difference was smaller than in the main analysis because most of the 73 004 composite events were losses to follow-up, but the absolute difference was unchanged, so a larger share of rivastigmine users leaving the data, through death or dropout, does not explain the main result. Both formulations of rivastigmine were associated with a lower risk of add-on at 24 months, the transdermal patch (risk difference *−*4.5 per 100; risk ratio 0.54) more than the oral forms (*−*2.7 per 100; 0.71; 8 110 users). The difference was present in every age group and became somewhat larger with age: the 24-month risk ratios were 0.64 in patients younger than 75 years, 0.57 at 75–84 years and 0.53 at 85 years or older.

The E-value for the 24-month risk ratio of rivastigmine versus donepezil was 2.84 (2.65 for the upper confidence limit): an unmeasured confounder associated with both the choice of rivastigmine and memantine add-on by a risk ratio of 2.84 each, beyond the measured covariates, could explain away the estimate, but weaker confounding could not. For galantamine versus donepezil the E-value was 1.36 (1.20 for the confidence limit).

The data quality checks are summarized in Supplementary Figure S1. Late processing affected 2.5% of records (1 to 3 months), and exact duplicate rows 0.05%. Within-year monthly retention of identifiers had a median of 91.5% across state-years (range 70.3% to 98.9%); 9 of 215 state-years were below 80%, mostly in Rio Grande do Sul. Of 185 December-to-January transitions, 16 fell more than 10 percentage points below the state’s within-year mean, most coinciding with months of low record volume. The proportion of patients with a record in one year who also had a record in the next was stable (75.4% to 80.8% in every year from 2018 to 2024), which argues against a yearly change of identifier encoding. A death code was recorded for 0.31% to 0.47% of treated patients per year nationally, and never in five states (Bahia, Goiás, Maranhão, Minas Gerais and Rio Grande do Sul).

## 4 Discussion

### 4.1 Key results

In this nationwide cohort of 142 758 patients starting a ChEI in the Brazilian public health system, starting rivastigmine was associated with a lower risk of memantine add-on over four years than starting donepezil or galantamine, with an absolute difference of about 4 per 100 patients at 2 years and 7 per 100 at 4 years relative to donepezil. The difference between galantamine and donepezil was small and depended on adjustment. The results were robust to weight truncation, to the censoring model, to the handling of a data quality problem affecting two large states, to counting deaths and dropouts as events, and across age groups and both formulations of rivastigmine. The dispensing data were suitable for following patients over time, but not for ascertaining deaths.

### 4.2 Limitations

The emulation departs from the target trial in several ways (Table 1). Assignment was not random, so the comparison depends on conditional exchangeability given a small set of recorded covariates; eligibility could not be checked against cognitive and functional staging; time zero and follow-up were measured in calendar months; the outcome was a dispensing event used as a proxy of clinical progression; and deaths, which would be observed in a trial, were largely missing. The plausibility of the identifying assumptions is discussed below.

The most important limitation is that deaths were recorded for less than 0.5% of treated patients per year, far below the mortality expected in an older population with dementia. Death could not be analysed as an outcome or as a competing event, and the analysis estimates the risk of add-on in a hypothetical scenario without censoring, including censoring by death. Rivastigmine users were older, so unrecorded deaths are probably more frequent among them; if deaths occur preferentially in patients who would otherwise have progressed, and are not predicted by the recorded covariates, the censoring weights would not remove the bias, and the lower risk of add-on with rivastigmine would be overestimated. Two additional analyses argue against a large bias of this kind, although they cannot exclude it: the difference was unchanged when deaths and dropouts were counted as events, and it was present in patients younger than 75 years, in whom deaths are uncommon. The slightly larger difference at older ages is, however, compatible with some contribution of unrecorded deaths.

Disease severity, education, comorbidities and co-medications are not recorded, a limitation of DATASUS administrative records for observational research that we have described previously [38]. If clinicians prefer rivastigmine for patients at an earlier or more stable stage, residual confounding would also favour rivastigmine; if they prefer it for frailer patients who are less likely to be considered for combination therapy, the effect would be in the same direction. We could not measure these biases directly; the E-value of 2.84 indicates that an unmeasured factor would need to be strongly associated with both drug choice and memantine add-on to explain away the difference between rivastigmine and donepezil. Access to specialists who can prescribe and the availability of each drug in state pharmacies may also influence which ChEI is started; the analysis was adjusted for state, but local access and stock shortages were not measured. Dispensing does not guarantee use. Diagnostic codes may not reflect the clinical subtype: 53.8% of patients were coded as early-onset disease (G30.0), although the mean age was 77 years. The outcome is a proxy of progression that depends on the organization of care and on reassessments that are not always performed [12]. Follow-up was relatively short (median 14 months), and many patients were lost to follow-up, although the censoring model and sensitivity analyses gave consistent results. Retention checks showed irregular dispensing and low-volume months in some states, and 29 monthly files were unavailable; we treated these months as unobserved but could not correct months with incomplete files. Switching between ChEIs was ignored, so estimates refer to the initial choice of drug.

### 4.3 Interpretation

Randomized trials have not shown differences in efficacy between the three ChEIs [4, 7], and the size of the rivastigmine difference is larger than would be expected from the trial evidence. Several explanations other than slower disease progression are plausible, and the data do not allow us to separate them. Memantine add-on depends on a clinical reassessment and a new authorization, so the outcome reflects prescribing and access as well as progression; if patients started on rivastigmine are followed by different prescribers or services, or reassessed less often, add-on would be less frequent without any difference in disease course. Most rivastigmine users started with the transdermal patch, which may be preferred for older or frailer patients and for those with swallowing difficulties or poor oral adherence; such patients may be less likely to be considered for combination therapy. Unmeasured severity, discussed above, would act in the same direction. The difference was smaller with oral than with transdermal rivastigmine, so part of it may relate to the patch itself, for example through better persistence, or to the patients for whom it is chosen.

In the Swedish Dementia Registry, ChEI users had slightly slower cognitive decline and a 27% lower risk of death than matched non-users over an average of 5 years, and galantamine was the only ChEI associated with a lower risk of severe dementia [8]. We did not observe an advantage for galantamine, and the lower risk of add-on with rivastigmine has no counterpart in that study; the outcomes, however, differ (severe dementia defined by cognitive score versus a prescribing event). Registries of this kind record cognitive scores and deaths, which the Brazilian dispensing data lack. Our findings are therefore best seen as a signal that deserves confirmation with linked mortality data and clinical measures, rather than as evidence that one drug slows progression. The analysis also showed that the design problems of administrative data can be large: without the detection of the shared health card number, thousands of patients in two states would have appeared to be lost to follow-up in a single month.

### 4.4 Generalisability

The cohort includes all patients who started a ChEI through the public specialized pharmaceutical component, which is the main source of these drugs for most of the Brazilian population. The results apply to patients who meet the protocol criteria and obtain their medicines from the SUS, and may not apply to patients treated privately or outside the protocol. Coverage differed markedly between states: São Paulo and Paraná contributed almost two thirds of the cohort, while several states in the North and Northeast contributed few patients. The comparison between drugs was adjusted for state, but the absolute risks reflect the mix of services in the states with the largest numbers of patients.

Future work should link the dispensing records to the Mortality Information System and to clinical data with cognitive staging, which would allow death to be modelled as a competing event and severity to be included in the adjustment. Comparisons of the oral and transdermal formulations of rivastigmine, and analyses of reassessment practices by service, would help distinguish effects of the drug from effects of the pathway of care, as would comparisons between municipalities with different coverage of primary care by the Family Health Strategy, which may affect when treatment starts and how often patients are reassessed. Linking dispensing records to the Mortality Information System, or recording the exit reason when dispensing stops, would make these data suitable for studying mortality. A temporal validation in patients who start treatment from January 2026 has been registered prospectively, with the analysis code fixed before any of its data were accessed [39].

## 5 Conclusions

In routine SUS care, patients who started rivastigmine had a lower risk of memantine add-on than those who started donepezil or galantamine, while galantamine and donepezil were similar. Because the dispensing data do not record disease severity or death reliably, these differences cannot be attributed to slower progression without confirmation in linked data. Nationwide SUS dispensing records can support target trial emulation when their quality is checked and data problems are handled explicitly.

## Declarations

### Registration

The protocol of this study was not registered. A temporal validation in patients starting treatment from January 2026 was registered prospectively, before any of its data were accessed (OSF Registries, doi:10.17605/OSF.IO/AXPN5).

### Author contributions

H.R.F.: conceptualization, methodology, software, data curation, formal analysis, visualization, writing (original draft), and writing (review and editing). G.I.C.: conceptualization, validation, and writing (review and editing). P.V.F.S., J.P.Q. and M.B.S.A.: validation, and writing (review and editing). All authors read and approved the final manuscript.

### Use of AI-assisted tools

Claude Opus 5.5 (Anthropic) was used to help write the analysis code and to draft and edit the manuscript. H.R.F. designed the study, reviewed and ran all code, and checked the results and the text; all authors reviewed the manuscript and take full responsibility for its content.

## Data Availability

All data produced are available online at DATASUS through the microdatasus CRAN package (https://cran.r-project.org/web/packages/microdatasus/index.html). The analysis code (download, pseudonymization, quality checks, cohort construction and IPTW analysis) is available at osf.io/a8yhw. Pseudonymized patient-level data are not shared.

https://osf.io/a8yhw/overview

https://cran.r-project.org/web/packages/microdatasus/index.html

## Supplementary material

**Figure S1:**
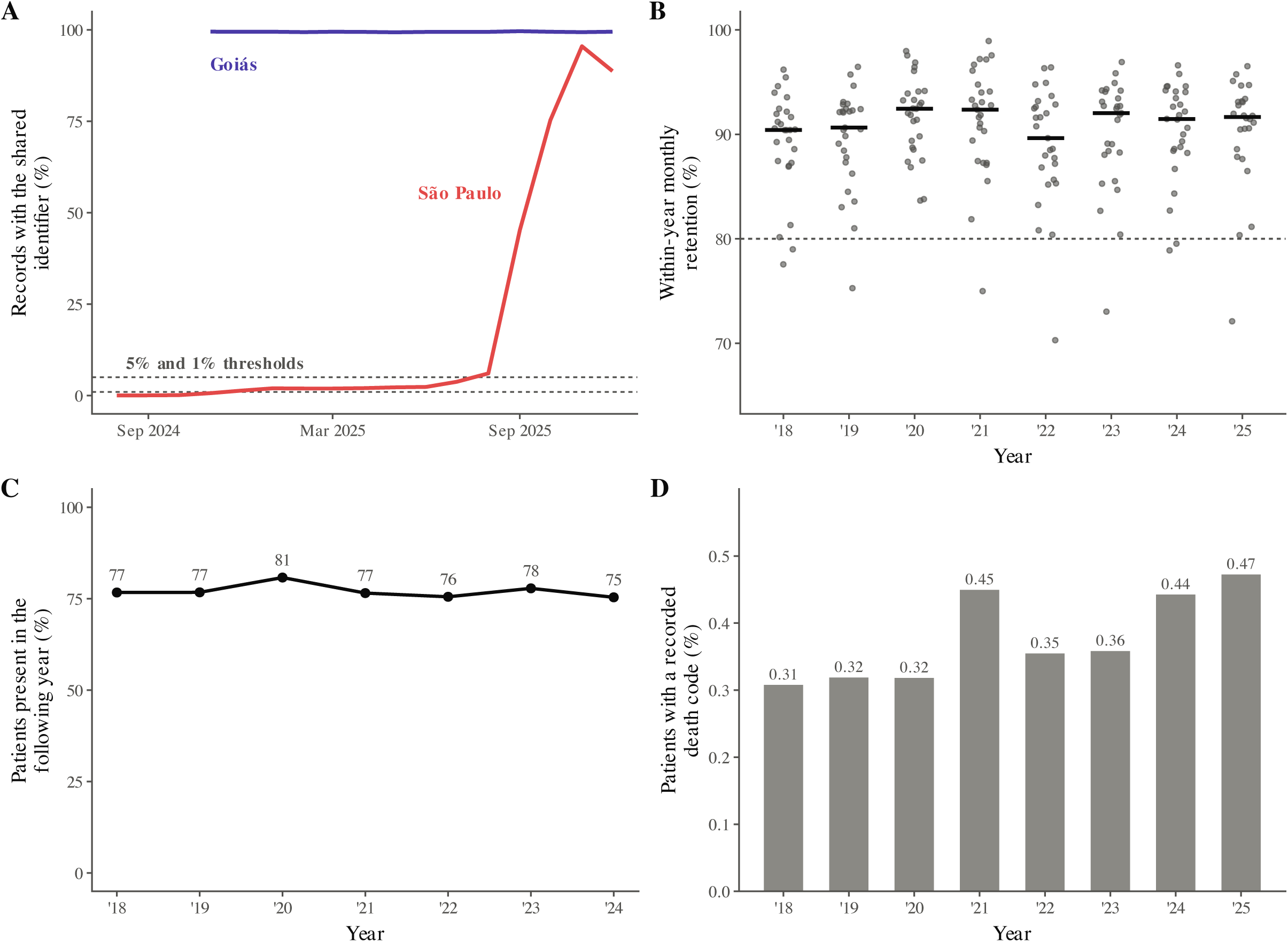
Data quality. (A) Share of records carrying the shared (placeholder) health card number in Goiás and São Paulo, by competence month; dotted lines mark the 1% and 5% thresholds. (B) Withinyear mean monthly retention of identifiers in each state and year (dots) and median across states (bars); dotted line, 80%. (C) Proportion of patients with a record in one year who also had a record in the following year. (D) Proportion of treated patients with a recorded death code per year.

**Table S1:** STROBE checklist for cohort studies.

| Section | Item | Recommendation | Location in the manuscript |
|---|---|---|---|
| Title and abstract | 1a | Indicate the study’s design with a commonly used term in the title or the abstract | Title; Abstract (Methods) |
|  | 1b | Provide in the abstract an informative and balanced summary of what was done and what was found | Abstract |
| Introduction: back-ground/rationale | 2 | Explain the scientific background and rationale for the investigation | Introduction, Background |
| Objectives | 3 | State specific objectives, including any prespecified hypotheses | Introduction, Objectives |
| Methods: study design | 4 | Present key elements of study design early in the paper | Methods, Study design; Table 1 |
| Setting | 5 | Describe the setting, locations, and relevant dates, including periods of recruitment, exposure, follow-up, and data collection | Methods, Setting |
| Participants | 6a | Give the eligibility criteria, and the sources and methods of selection of participants; describe methods of follow-up | Methods, Participants; Outcome and follow-up |
|  | 6b | For matched studies, give matching criteria and number of exposed and unexposed | Not applicable (no matching) |
| Variables | 7 | Clearly define all outcomes, exposures, predictors, potential confounders, and effect modifiers; give diagnostic criteria, if applicable | Methods, Variables |
| Data sources and measurement | 8 | For each variable of interest, give sources of data and details of methods of assessment; describe comparability of assessment methods if there is more than one group | Methods, Data sources and measurement |
| Bias | 9 | Describe any efforts to address potential sources of bias | Methods, Bias; Data quality assessment and handling |
| Study size | 10 | Explain how the study size was arrived at | Methods, Study size |
| Quantitative variables | 11 | Explain how quantitative variables were handled in the analyses; if applicable, describe which groupings were chosen and why | Methods, Quantitative variables |
| Statistical methods | 12a | Describe all statistical methods, including those used to control for confounding | Methods, Statistical methods (including estimands and identifying assumptions) |
|  | 12b | Describe any methods used to examine subgroups and interactions | Methods, Statistical methods (Subgroups and interactions; Additional analyses) |
|  | 12c | Explain how missing data were addressed | Methods, Statistical methods |
|  | 12d | If applicable, explain how loss to follow-up was addressed | Methods, Statistical methods; Outcome and follow-up |
|  | 12e | Describe any sensitivity analyses | Methods, Statistical methods |
| Results: participants | 13a | Report numbers of individuals at each stage of study | Results, Participants; Figure 1 |
|  | 13b | Give reasons for non-participation at each stage | Figure 1 |
|  | 13c | Consider use of a flow diagram | Figure 1 |
| Descriptive data | 14a | Give characteristics of study participants and information on exposures and potential confounders | Table 2 |
|  | 14b | Indicate number of participants with missing data for each variable of interest | Table 2 and footnote; Methods (missing data) |
|  | 14c | Summarise follow-up time (e.g., average and total amount) | Results, Descriptive data; Table 2 |
| Outcome data | 15 | Report numbers of outcome events or summary measures over time | Results, Outcome data; Table 3; Figure 2A |
| Main results | 16a | Give unadjusted estimates and, if applicable, confounder-adjusted estimates and their precision; make clear which confounders were adjusted for and why they were included | Results, Main results; Table 3; Figure 2 |
|  | 16b | Report category boundaries when continuous variables were categorized | Methods, Quantitative variables; Table 2 |
|  | 16c | If relevant, consider translating estimates of relative risk into absolute risk for a meaningful time period | Table 3 (risk differences per 100 patients at 12–48 months) |
| Other analyses | 17 | Report other analyses done, e.g., analyses of subgroups and interactions, and sensitivity analyses | Results, Other analyses; Figure 3B; Tables S4, S20 and S21; Figure S1 |
| Discussion: key results | 18 | Summarise key results with reference to study objectives | Discussion, Key results |
| Limitations | 19 | Discuss limitations of the study, taking into account sources of potential bias or imprecision; discuss both direction and magnitude of any potential bias | Discussion, Limitations |
| Interpretation | 20 | Give a cautious overall interpretation of results considering objectives, limitations, multiplicity of analyses, results from similar studies, and other relevant evidence | Discussion, Interpretation |
| Generalisability | 21 | Discuss the generalisability (external validity) of the study results | Discussion, Generalisability |
| Other information: funding | 22 | Give the source of funding and the role of the funders for the present study and, if applicable, for the original study on which the present article is based | Declarations, Funding |

**Table S2:**
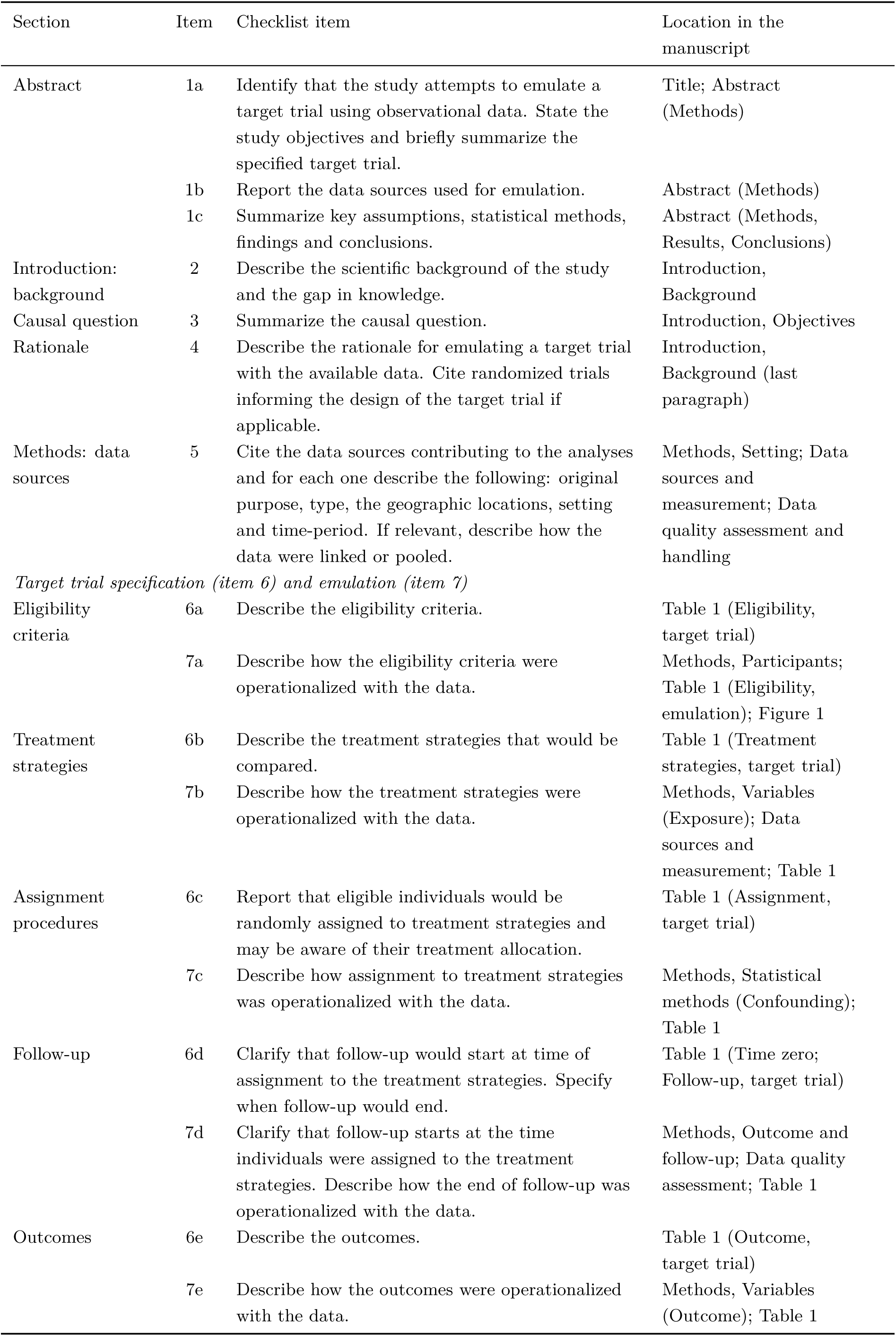
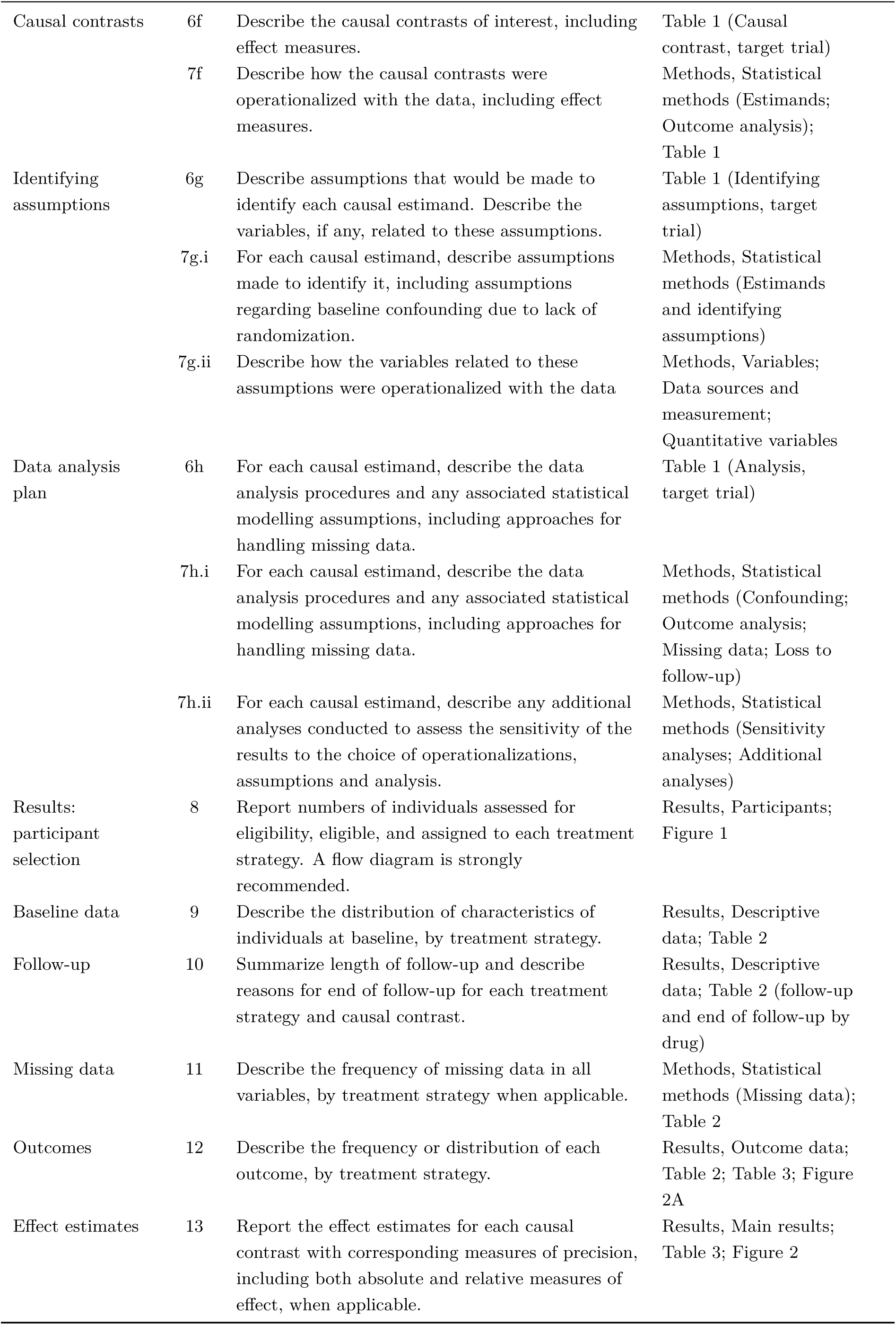
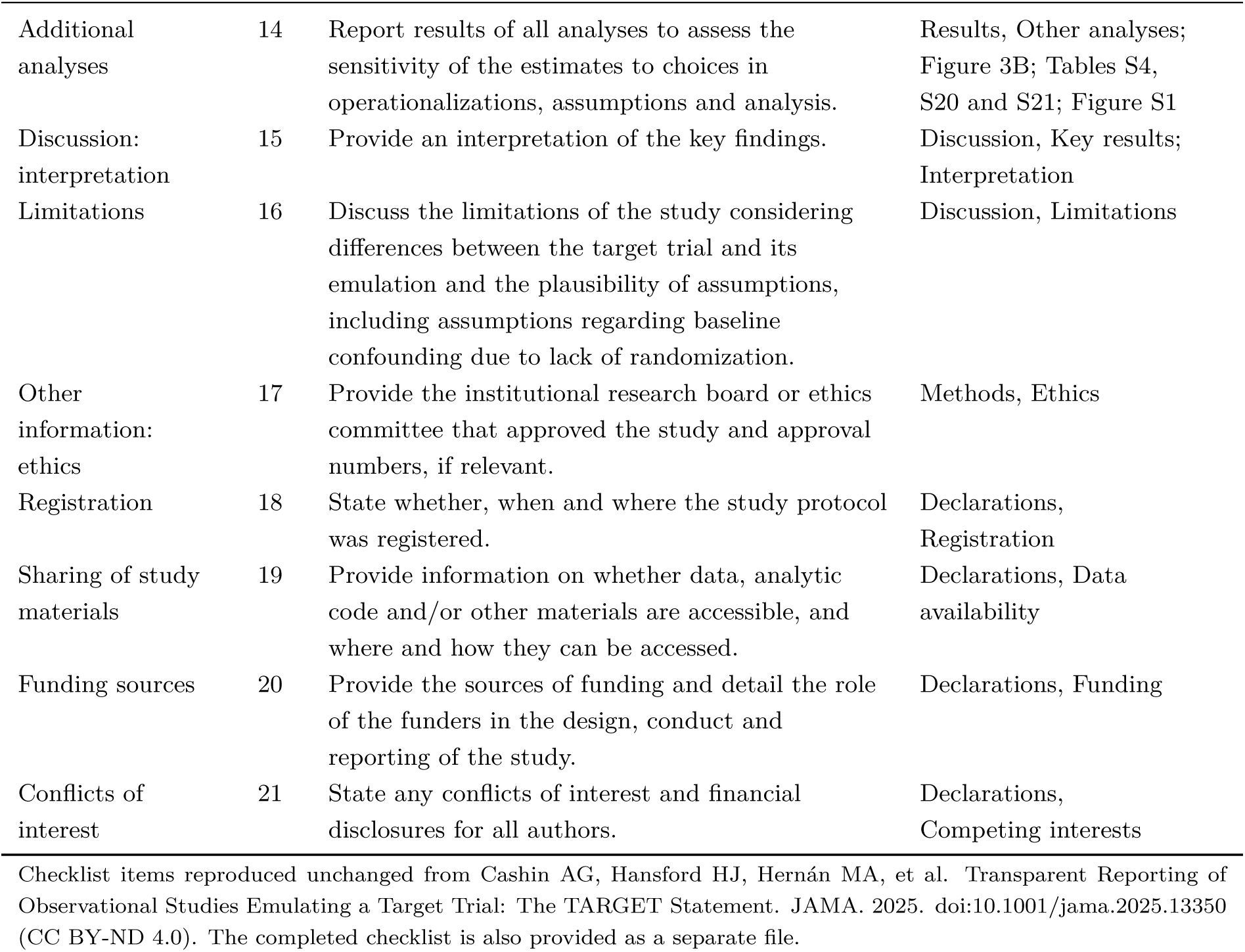
TARGET checklist for observational studies emulating a target trial [19].

| Section | Item | Checklist item | Location in the manuscript |
|---|---|---|---|
| Abstract | 1a | Identify that the study attempts to emulate a target trial using observational data. State the study objectives and briefly summarize the specified target trial. | Title; Abstract (Methods) |
|  | 1b | Report the data sources used for emulation. | Abstract (Methods) |
|  | 1c | Summarize key assumptions, statistical methods, findings and conclusions. | Abstract (Methods, Results, Conclusions) |
| Introduction: background | 2 | Describe the scientific background of the study and the gap in knowledge. | Introduction, Background |
| Causal question | 3 | Summarize the causal question. | Introduction, Objectives |
| Rationale | 4 | Describe the rationale for emulating a target trial with the available data. Cite randomized trials informing the design of the target trial if applicable. | Introduction, Background (last paragraph) |
| Methods: data sources | 5 | Cite the data sources contributing to the analyses and for each one describe the following: original purpose, type, the geographic locations, setting and time-period. If relevant, describe how the data were linked or pooled. | Methods, Setting; Data sources and measurement; Data quality assessment and handling |
| *Target trial specification (item 6) and emulation (item 7)* |  |  |  |
| Eligibility criteria | 6a | Describe the eligibility criteria. | Table 1 (Eligibility, target trial) |
|  | 7a | Describe how the eligibility criteria were operationalized with the data. | Methods, Participants; Table 1 (Eligibility, emulation); Figure 1 |
| Treatment strategies | 6b | Describe the treatment strategies that would be compared. | Table 1 (Treatment strategies, target trial) |
|  | 7b | Describe how the treatment strategies were operationalized with the data. | Methods, Variables (Exposure); Data sources and measurement; Table 1 |
| Assignment procedures | 6c | Report that eligible individuals would be randomly assigned to treatment strategies and may be aware of their treatment allocation. | Table 1 (Assignment, target trial) |
|  | 7c | Describe how assignment to treatment strategies was operationalized with the data. | Methods, Statistical methods (Confounding); Table 1 |
| Follow-up | 6d | Clarify that follow-up would start at time of assignment to the treatment strategies. Specify when follow-up would end. | Table 1 (Time zero; Follow-up, target trial) |
|  | 7d | Clarify that follow-up starts at the time individuals were assigned to the treatment strategies. Describe how the end of follow-up was operationalized with the data. | Methods, Outcome and follow-up; Data quality assessment; Table 1 |
| Outcomes | 6e | Describe the outcomes. | Table 1 (Outcome, target trial) |
|  | 7e | Describe how the outcomes were operationalized with the data. | Methods, Variables (Outcome); Table 1 |
| Causal contrasts | 6f | Describe the causal contrasts of interest, including effect measures. | Table 1 (Causal contrast, target trial) |
|  | 7f | Describe how the causal contrasts were operationalized with the data, including effect measures. | Methods, Statistical methods (Estimands; Outcome analysis); Table 1 |
| Identifying assumptions | 6g | Describe assumptions that would be made to identify each causal estimand. Describe the variables, if any, related to these assumptions. | Table 1 (Identifying assumptions, target trial) |
|  | 7g.i | For each causal estimand, describe assumptions made to identify it, including assumptions regarding baseline confounding due to lack of randomization. | Methods, Statistical methods (Estimands and identifying assumptions) |
|  | 7g.ii | Describe how the variables related to these assumptions were operationalized with the data | Methods, Variables; Data sources and measurement; Quantitative variables |
| Data analysis plan | 6h | For each causal estimand, describe the data analysis procedures and any associated statistical modelling assumptions, including approaches for handling missing data. | Table 1 (Analysis, target trial) |
|  | 7h.i | For each causal estimand, describe the data analysis procedures and any associated statistical modelling assumptions, including approaches for handling missing data. | Methods, Statistical methods (Confounding; Outcome analysis; Missing data; Loss to follow-up) |
|  | 7h.ii | For each causal estimand, describe any additional analyses conducted to assess the sensitivity of the results to the choice of operationalizations, assumptions and analysis. | Methods, Statistical methods (Sensitivity analyses; Additional analyses) |
| Results: participant selection | 8 | Report numbers of individuals assessed for eligibility, eligible, and assigned to each treatment strategy. A flow diagram is strongly recommended. | Results, Participants; Figure 1 |
| Baseline data | 9 | Describe the distribution of characteristics of individuals at baseline, by treatment strategy. | Results, Descriptive data; Table 2 |
| Follow-up | 10 | Summarize length of follow-up and describe reasons for end of follow-up for each treatment strategy and causal contrast. | Results, Descriptive data; Table 2 (follow-up and end of follow-up by drug) |
| Missing data | 11 | Describe the frequency of missing data in all variables, by treatment strategy when applicable. | Methods, Statistical methods (Missing data); Table 2 |
| Outcomes | 12 | Describe the frequency or distribution of each outcome, by treatment strategy. | Results, Outcome data; Table 2; Table 3; Figure 2A |
| Effect estimates | 13 | Report the effect estimates for each causal contrast with corresponding measures of precision, including both absolute and relative measures of effect, when applicable. | Results, Main results; Table 3; Figure 2 |
| Additional analyses | 14 | Report results of all analyses to assess the sensitivity of the estimates to choices in operationalizations, assumptions and analysis. | Results, Other analyses; Figure 3B; Tables S4, S20 and S21; Figure S1 |
| Discussion: interpretation | 15 | Provide an interpretation of the key findings. | Discussion, Key results; Interpretation |
| Limitations | 16 | Discuss the limitations of the study considering differences between the target trial and its emulation and the plausibility of assumptions, including assumptions regarding baseline confounding due to lack of randomization. | Discussion, Limitations |
| Other information: ethics | 17 | Provide the institutional research board or ethics committee that approved the study and approval numbers, if relevant. | Methods, Ethics |
| Registration | 18 | State whether, when and where the study protocol was registered. | Declarations, Registration |
| Sharing of study materials | 19 | Provide information on whether data, analytic code and/or other materials are accessible, and where and how they can be accessed. | Declarations, Data availability |
| Funding sources | 20 | Provide the sources of funding and detail the role of the funders in the design, conduct and reporting of the study. | Declarations, Funding |
| Conflicts of interest | 21 | State any conflicts of interest and financial disclosures for all authors. | Declarations, Competing interests |

Checklist items reproduced unchanged from Cashin AG, Hansford HJ, Hernán MA, et al. Transparent Reporting of Observational Studies Emulating a Target Trial: The TARGET Statement. JAMA. 2025. doi:10.1001/jama.2025.13350 (CC BY-ND 4.0). The completed checklist is also provided as a separate file.

**Table S3:** Proportion of patient-months with an initial APAC authorization, by months since the index month (cohort members).

| Months since index | Patients with a dispensing | Initial APAC (%) |
|---|---|---|
| 0 | 142 758 | 100.0 |
| 1 | 136 031 | 9.5 |
| 2 | 132 079 | 9.6 |
| 3 | 113 489 | 97.7 |
| 4 | 114 098 | 14.2 |
| 5 | 111 879 | 12.4 |
| 6 | 80 308 | 90.0 |
| 7 | 87 664 | 26.8 |
| 8 | 88 672 | 18.4 |
| 9 | 82 691 | 75.7 |
| 10 | 81 772 | 27.8 |
| 11 | 80 115 | 19.8 |
| 12 | 68 484 | 69.6 |

Initial authorizations recur every three months (quarterly renewal), so the APAC type does not identify new users.

**Table S4:** Sensitivity analyses: risk differences (per 100 patients) and risk ratios at 24 and 36 months.

|  | GAL vs DON |  | RIV vs DON |  | GAL vs RIV |  |
|---|---|---|---|---|---|---|
| Analysis | RD | RR | RD | RR | RD | RR |
| *24 months* |  |  |  |  |  |  |
| Main analysis | −0.7 | 0.93 | −4.1 | 0.58 | 3.4 | 1.60 |
| Untruncated weights | −0.7 | 0.93 | −4.1 | 0.58 | 3.4 | 1.61 |
| Without censoring weights | −0.7 | 0.93 | −4.2 | 0.57 | 3.5 | 1.61 |
| Unweighted | 0.6 | 1.07 | −3.0 | 0.67 | 3.6 | 1.60 |
| Placeholder threshold 1% | −0.7 | 0.93 | −4.4 | 0.57 | 3.6 | 1.62 |
| *36 months* |  |  |  |  |  |  |
| Main analysis | −1.0 | 0.93 | −5.5 | 0.61 | 4.5 | 1.53 |
| Untruncated weights | −1.0 | 0.93 | −5.5 | 0.61 | 4.5 | 1.53 |
| Without censoring weights | −1.0 | 0.93 | −5.7 | 0.60 | 4.7 | 1.55 |
| Unweighted | 0.6 | 1.05 | −4.3 | 0.67 | 5.0 | 1.56 |
| Placeholder threshold 1% | −1.0 | 0.94 | −6.0 | 0.60 | 5.0 | 1.56 |

DON, donepezil; GAL, galantamine; RIV, rivastigmine; RD, risk difference; RR, risk ratio.

## Data quality checks

**Table S5:** Prespecified data quality criteria and their assessment in the national data.

| Criterion | Assessment | Result |
|---|---|---|
| Within-year monthly retention of identifiers ≥80% in every state-year | Not met | 9 of 215 state-years below 80% |
| December–January retention not more than 10 percentage points below the within-year mean | Not met | 16 of 185 transitions |
| No yearly change of identifier encoding (year-to-year carry-over ≥50%) | Met | 75.4% to 80.8% |
| Placeholder identifiers detected and handled | Met (records excluded; state-specific end of data) | 1 identifier |
| ≥200 new users per drug | Met | 74 165 / 30 691 / 37 902 |
| ≥30 memantine add-on events per drug | Met | 5 479 / 2 391 / 1 942 |
| Each drug ≥5% of new users in every state (positivity) | Met | All states |
| Age and sex ≥95% complete | Met | 100% and 100% |
| Recording of deaths plausible | Not met | At most 0.47% of patients per year |

Criteria not met did not stop the analysis: retention problems were handled by treating unavailable files as unobserved months, and the poor recording of deaths is addressed in the analysis (censoring weights) and in the Discussion.

**Table S6:** Monthly SIA/SUS medication files (AM) unavailable on DATASUS, January 2018 to December 2025 (29 of 2 592 state-months).

| State | Files | Competence months |
|---|---|---|
| Acre | 4 | 2018-08, 2018-10, 2022-06, 2022-09 |
| Amazonas | 3 | 2020-12, 2022-01, 2024-10 |
| Amapá | 5 | 2020-05, 2023-03, 2024-09, 2024-10, 2025-06 |
| Bahia | 1 | 2020-07 |
| Maranhão | 1 | 2023-04 |
| Pará | 1 | 2019-04 |
| Paraíba | 2 | 2018-12, 2023-07 |
| Rondônia | 4 | 2019-09, 2019-11, 2021-02, 2021-06 |
| Rio Grande do Sul | 7 | 2020-11, 2021-04, 2022-06, 2022-07, 2022-08, 2024-04, 2024-05 |
| Tocantins | 1 | 2025-05 |
| Total | 29 |  |

These months were treated as unobserved: a gap in dispensing that overlapped an unavailable file did not count towards the 3-month limit that defined loss to follow-up.

**Table S7:** Records by processing year: late processing and duplicate records.

| Year | Records | Late (%) | Lag 1 mo | Lag 2 mo | Lag 3 mo | Exact duplicates | Repeated (%) |
|---|---|---|---|---|---|---|---|
| 2018 | 938 441 | 3.6 | 11 819 | 4 672 | 17 193 | 1 793 | 0.79 |
| 2019 | 945 800 | 3.9 | 24 676 | 7 029 | 5 526 | 1 022 | 0.30 |
| 2020 | 952 636 | 2.0 | 14 575 | 3 140 | 1 060 | 553 | 0.45 |
| 2021 | 974 015 | 2.2 | 14 107 | 5 247 | 1 991 | 258 | 0.44 |
| 2022 | 927 696 | 3.0 | 18 864 | 6 986 | 2 245 | 156 | 0.53 |
| 2023 | 972 773 | 1.9 | 12 806 | 3 046 | 2 801 | 38 | 0.40 |
| 2024 | 1 012 114 | 2.1 | 15 106 | 5 943 | 601 | 332 | 0.37 |
| 2025 | 1 061 788 | 1.5 | 12 919 | 2 777 | 654 | 0 | 0.30 |

Late: records whose competence month preceded the processing month (lag in months); no record had a lag greater than 3 months or a competence month later than the processing month. Exact duplicates: identical rows, removed. Repeated: more than one record of the same drug for the same patient and month, as a percentage of records; these were collapsed into one person-month.

**Table S8:** Completeness of baseline covariates in the study cohort, by drug (%).

| Variable | Donepezil | Galantamine | Rivastigmine |
|---|---|---|---|
| Age | 100.0 | 100.0 | 100.0 |
| Sex | 100.0 | 100.0 | 100.0 |
| ICD-10 code | 100.0 | 100.0 | 100.0 |
| Race/colour informed | 77.3 | 78.9 | 75.3 |
| Weight and height plausible (BMI) | 97.6 | 97.1 | 97.0 |

In the full set of dispensing records (2017–2025), none of the 20 fields used (competence month, procedure, authorization, APAC type, dates, ICD-10 code, age, sex, race/colour, municipality and state, establishment, exit reason, death indicator, weight and height) had missing values, except the date of the event (AP\_DTOCOR), which was empty in 98% to 99.7% of records per year and was not used. Race/colour “not informed” is a valid code and was analysed as a separate category.

**Table S9:** Dispensing records by SIGTAP procedure code and year.

| Code | Medicine | 2018 | 2019 | 2020 | 2021 | 2022 | 2023 | 2024 | 2025 |
|---|---|---|---|---|---|---|---|---|---|
| 0604130015 | Donepezil 5 mg tablet | 194 638 | 168 880 | 156 146 | 151 836 | 138 417 | 124 965 | 121 619 | 117 208 |
| 0604130023 | Donepezil 10 mg tablet | 333 274 | 352 291 | 358 067 | 358 212 | 343 385 | 357 750 | 369 165 | 384 135 |
| 0604130031 | Galantamine 8 mg ER capsule | 35 980 | 36 591 | 36 865 | 40 103 | 40 725 | 42 071 | 45 820 | 49 574 |
| 0604130040 | Galantamine 16 mg ER capsule | 44 825 | 40 691 | 42 498 | 43 394 | 47 568 | 52 308 | 52 114 | 58 685 |
| 0604130058 | Galantamine 24 mg ER capsule | 83 824 | 82 338 | 79 235 | 79 599 | 79 159 | 91 696 | 78 869 | 92 567 |
| 0604130066 | Rivastigmine 1.5 mg capsule | 43 671 | 34 995 | 21 660 | 23 563 | 15 575 | 14 142 | 14 522 | 15 131 |
| 0604130074 | Rivastigmine 2 mg/mL oral solution | 15 531 | 15 266 | 13 112 | 9 155 | 5 173 | 2 632 | 1 958 | 1 495 |
| 0604130082 | Rivastigmine 3 mg capsule | 65 527 | 56 183 | 50 335 | 40 046 | 17 247 | 18 904 | 17 926 | 17 197 |
| 0604130090 | Rivastigmine 4.5 mg capsule | 35 549 | 28 823 | 24 119 | 19 072 | 9 104 | 8 693 | 7 272 | 7 221 |
| 0604130104 | Rivastigmine 6 mg capsule | 69 885 | 59 643 | 43 023 | 38 295 | 18 707 | 18 673 | 16 673 | 10 657 |
| 0604130120 | Rivastigmine 9 mg patch | 5 935 | 22 204 | 18 311 | 16 199 | 16 063 | 12 142 | 10 853 | 13 250 |
| 0604130139 | Rivastigmine 18 mg patch | 10 738 | 44 861 | 88 578 | 111 992 | 129 539 | 145 952 | 169 352 | 199 252 |
| 0604540078 | Memantine 10 mg tablet | 0 | 1 092 | 18 541 | 43 974 | 65 511 | 83 322 | 104 670 | 94 406 |

ER, extended release. All procedure codes in the records mapped to one of these 13 codes. All codes were first recorded in October 2017, except the rivastigmine 18 mg and 9 mg patches (January and March 2018) and memantine (September 2019). 5 084 records with competence months in 2017 (in files of 2018) are not shown.

**Table S10:** Patients with dispensing records by ICD-10 code of the main diagnosis and year.

| ICD-10 | 2018 | 2019 | 2020 | 2021 | 2022 | 2023 | 2024 | 2025 |
|---|---|---|---|---|---|---|---|---|
| F00.0 | 680 | 1 313 | 1 958 | 2 815 | 3 678 | 4 495 | 5 219 | 5 518 |
| F00.1 | 859 | 1 804 | 2 503 | 3 409 | 4 379 | 5 314 | 5 975 | 6 131 |
| F00.2 | 236 | 570 | 799 | 1 174 | 1 631 | 2 136 | 2 608 | 2 716 |
| G30.0 | 75 243 | 71 826 | 66 652 | 67 980 | 67 413 | 68 062 | 68 781 | 64 806 |
| G30.1 | 29 392 | 30 419 | 28 691 | 29 554 | 29 655 | 29 911 | 30 707 | 30 069 |
| G30.8 | 9 795 | 9 815 | 9 200 | 9 482 | 9 666 | 10 399 | 11 204 | 11 111 |

A patient may appear under more than one code in the same year. No other G30 or F00 subcodes were present.

**Table S11:**
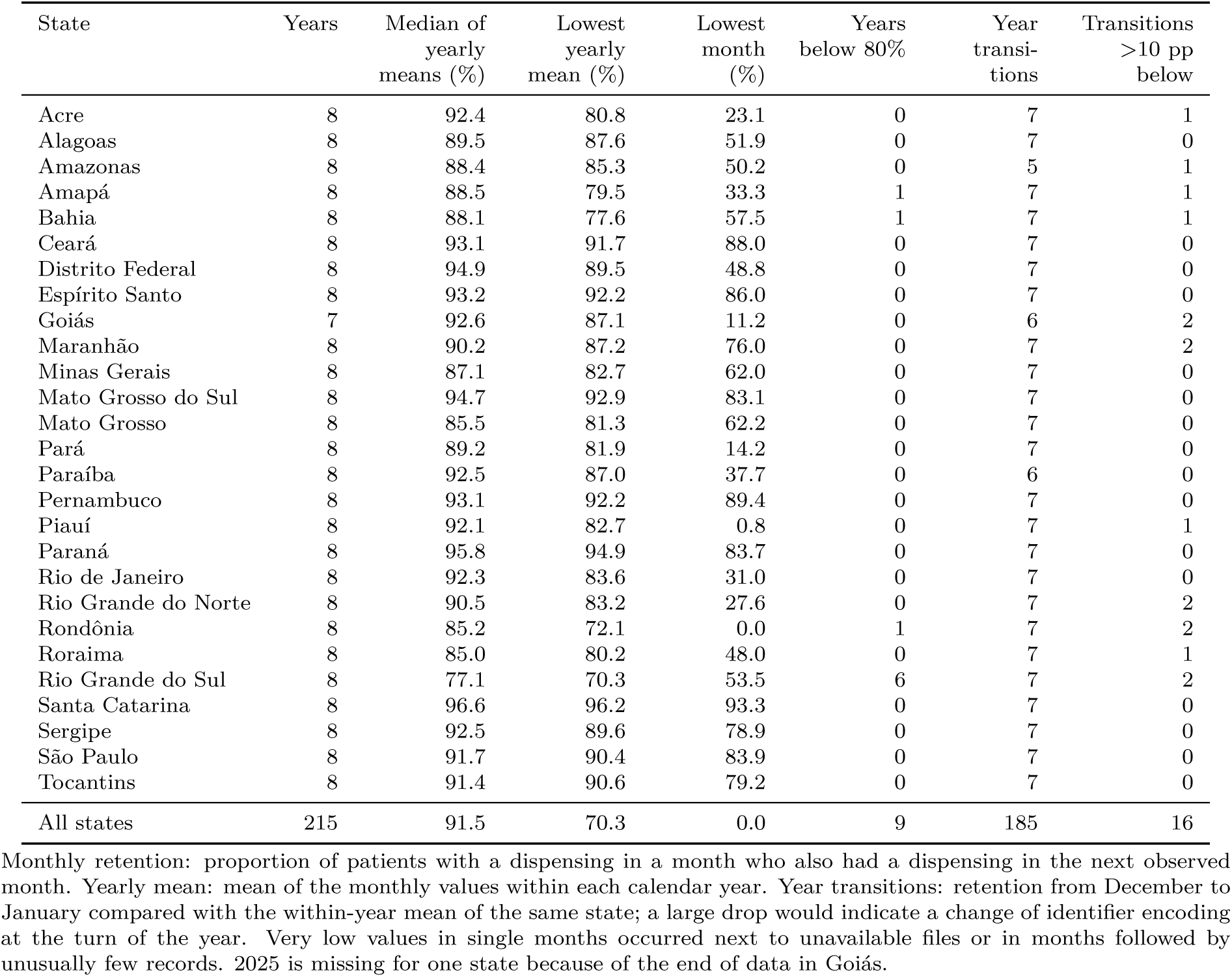
Monthly retention of pseudonymous identifiers by state, 2018–2025.

| State | Years | Median of yearly means (%) | Lowest yearly mean (%) | Lowest month (%) | Years below 80% | Year transitions | Transitions >10 pp below |
|---|---|---|---|---|---|---|---|
| Acre | 8 | 92.4 | 80.8 | 23.1 | 0 | 7 | 1 |
| Alagoas | 8 | 89.5 | 87.6 | 51.9 | 0 | 7 | 0 |
| Amazonas | 8 | 88.4 | 85.3 | 50.2 | 0 | 5 | 1 |
| Amapá | 8 | 88.5 | 79.5 | 33.3 | 1 | 7 | 1 |
| Bahia | 8 | 88.1 | 77.6 | 57.5 | 1 | 7 | 1 |
| Ceará | 8 | 93.1 | 91.7 | 88.0 | 0 | 7 | 0 |
| Distrito Federal | 8 | 94.9 | 89.5 | 48.8 | 0 | 7 | 0 |
| Espírito Santo | 8 | 93.2 | 92.2 | 86.0 | 0 | 7 | 0 |
| Goiás | 7 | 92.6 | 87.1 | 11.2 | 0 | 6 | 2 |
| Maranhão | 8 | 90.2 | 87.2 | 76.0 | 0 | 7 | 2 |
| Minas Gerais | 8 | 87.1 | 82.7 | 62.0 | 0 | 7 | 0 |
| Mato Grosso do Sul | 8 | 94.7 | 92.9 | 83.1 | 0 | 7 | 0 |
| Mato Grosso | 8 | 85.5 | 81.3 | 62.2 | 0 | 7 | 0 |
| Pará | 8 | 89.2 | 81.9 | 14.2 | 0 | 7 | 0 |
| Paraíba | 8 | 92.5 | 87.0 | 37.7 | 0 | 6 | 0 |
| Pernambuco | 8 | 93.1 | 92.2 | 89.4 | 0 | 7 | 0 |
| Piauí | 8 | 92.1 | 82.7 | 0.8 | 0 | 7 | 1 |
| Paraná | 8 | 95.8 | 94.9 | 83.7 | 0 | 7 | 0 |
| Rio de Janeiro | 8 | 92.3 | 83.6 | 31.0 | 0 | 7 | 0 |
| Rio Grande do Norte | 8 | 90.5 | 83.2 | 27.6 | 0 | 7 | 2 |
| Rondônia | 8 | 85.2 | 72.1 | 0.0 | 1 | 7 | 2 |
| Roraima | 8 | 85.0 | 80.2 | 48.0 | 0 | 7 | 1 |
| Rio Grande do Sul | 8 | 77.1 | 70.3 | 53.5 | 6 | 7 | 2 |
| Santa Catarina | 8 | 96.6 | 96.2 | 93.3 | 0 | 7 | 0 |
| Sergipe | 8 | 92.5 | 89.6 | 78.9 | 0 | 7 | 0 |
| São Paulo | 8 | 91.7 | 90.4 | 83.9 | 0 | 7 | 0 |
| Tocantins | 8 | 91.4 | 90.6 | 79.2 | 0 | 7 | 0 |
| All states | 215 | 91.5 | 70.3 | 0.0 | 9 | 185 | 16 |

Monthly retention: proportion of patients with a dispensing in a month who also had a dispensing in the next observed month. Yearly mean: mean of the monthly values within each calendar year. Year transitions: retention from December to January compared with the within-year mean of the same state; a large drop would indicate a change of identifier encoding at the turn of the year. Very low values in single months occurred next to unavailable files or in months followed by unusually few records. 2025 is missing for one state because of the end of data in Goiás.

**Table S12:**
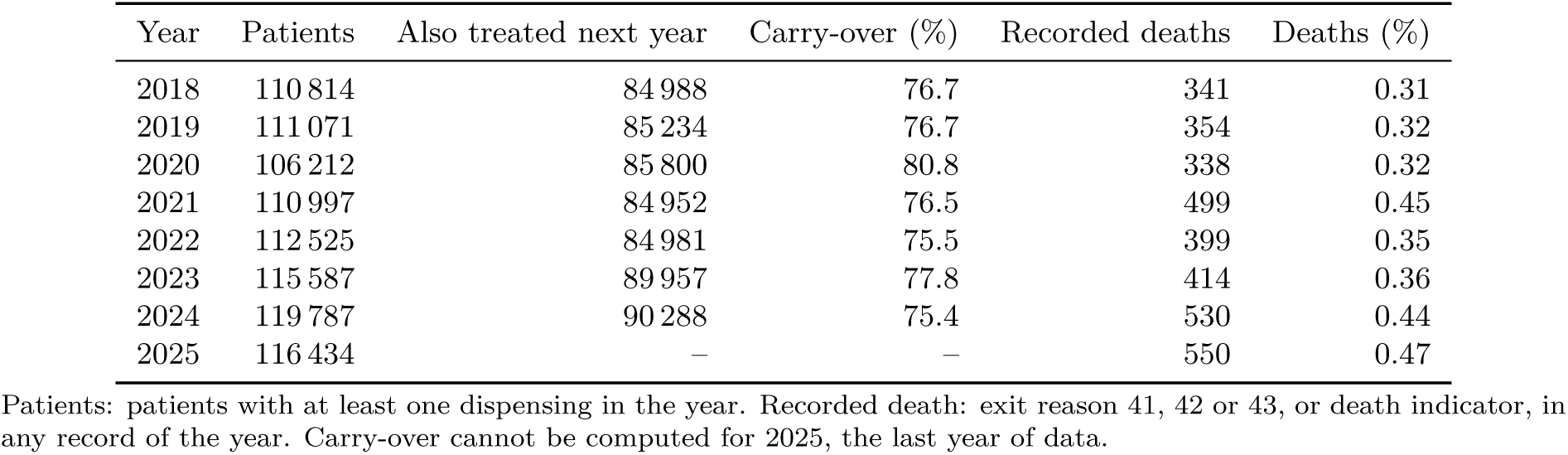
Year-to-year continuity of treated patients and recorded deaths, by year.

| Year | Patients | Also treated next year | Carry-over (%) | Recorded deaths | Deaths (%) |
|---|---|---|---|---|---|
| 2018 | 110 814 | 84 988 | 76.7 | 341 | 0.31 |
| 2019 | 111 071 | 85 234 | 76.7 | 354 | 0.32 |
| 2020 | 106 212 | 85 800 | 80.8 | 338 | 0.32 |
| 2021 | 110 997 | 84 952 | 76.5 | 499 | 0.45 |
| 2022 | 112 525 | 84 981 | 75.5 | 399 | 0.35 |
| 2023 | 115 587 | 89 957 | 77.8 | 414 | 0.36 |
| 2024 | 119 787 | 90 288 | 75.4 | 530 | 0.44 |
| 2025 | 116 434 | – | – | 550 | 0.47 |

Patients: patients with at least one dispensing in the year. Recorded death: exit reason 41, 42 or 43, or death indicator, in any record of the year. Carry-over cannot be computed for 2025, the last year of data.

**Table S13:** Dispensing records by exit reason (AP_MOTSAI) and death indicator (AP_OBITO), 2017– 2025.

| Exit reason (codes) | Records | % | Death indicator = 1 |
|---|---|---|---|
| Discharge (11–19) | 19 503 | 0.25 | 0 |
| Continued treatment (permanence) (21–28) | 7 747 029 | 99.51 | 0 |
| Transfer (31) | 5 379 | 0.07 | 0 |
| Death (41–43) | 3 435 | 0.04 | 3 435 |
| 41, certificate by attending physician | 3 027 | 0.04 | 3 027 |
| 42, certificate by forensic institute (IML) | 71 | 0.00 | 71 |
| 43, certificate by death verification service (SVO) | 337 | 0.00 | 337 |
| Administrative closure (51) | 9 917 | 0.13 | 0 |
| Total | 7 785 263 | 100.00 | 3 435 |

Codes and labels from the DATASUS TabWin reference table for the SIA exit and continuation reasons (retrieved with microdatasus); codes 11–18 are types of discharge, 21–28 reasons for continuing treatment, and 41–43 deaths by the service that issued the death certificate. The death indicator was set only in records with exit reasons 41–43.

**Table S14:**
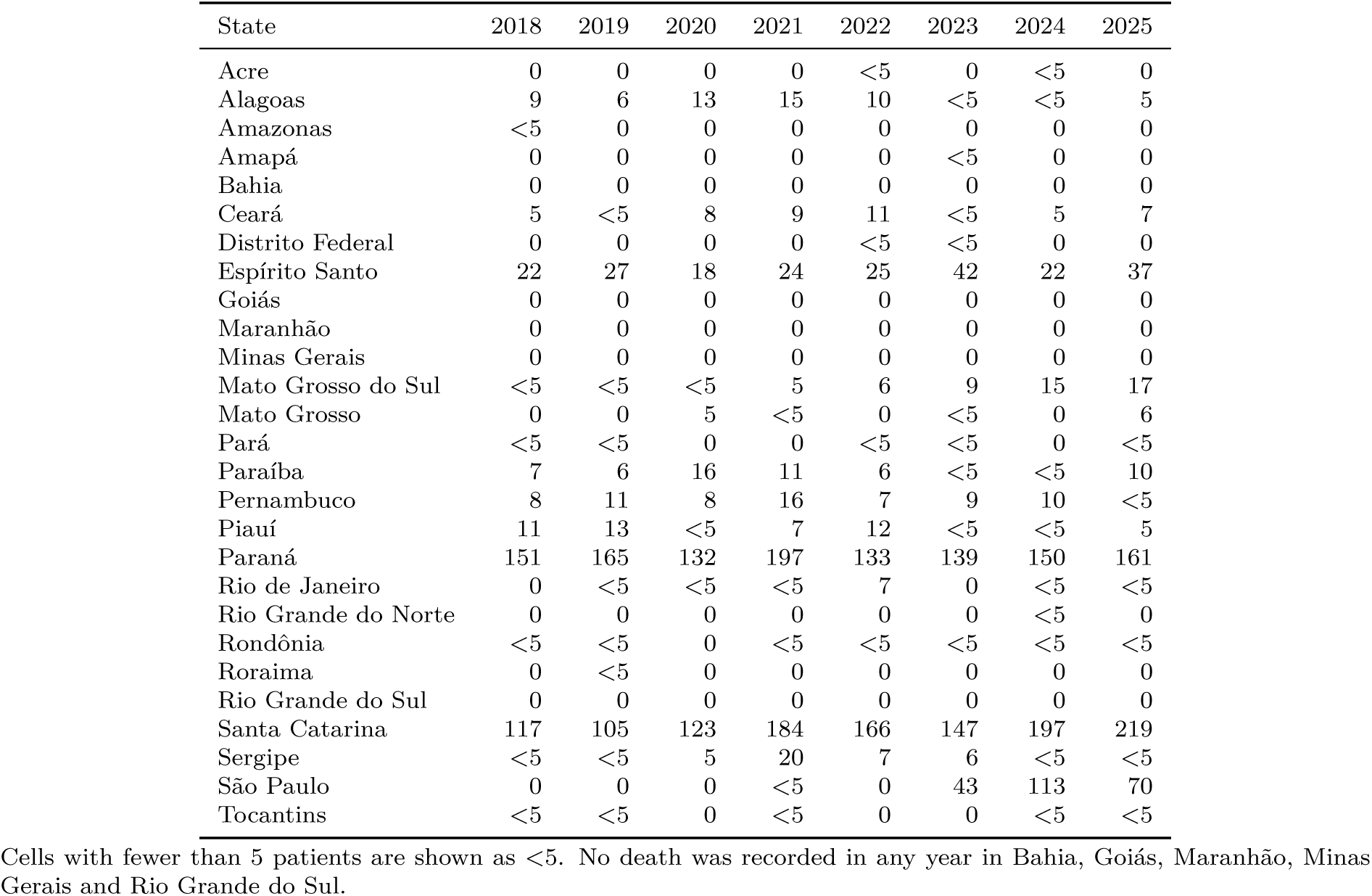
Patients with a recorded death by state and year.

**Table S15:** Share of records carrying the placeholder health card number in Goiás and São Paulo, by competence month.

| Goiás |  |  | São Paulo |  |  |
|---|---|---|---|---|---|
| Month | Records | Placeholder (%) | Month | Records | Placeholder (%) |
| 2024-11 | 1 593 | 99.5 | 2024-08 | 38 447 | 0.0 |
| 2024-12 | 1 682 | 99.5 | 2024-09 | 38 543 | 0.1 |
| 2025-01 | 1 710 | 99.5 | 2024-10 | 38 923 | 0.1 |
| 2025-02 | 1 738 | 99.4 | 2024-11 | 39 530 | 0.7 |
| 2025-03 | 1 703 | 99.5 | 2024-12 | 40 798 | 1.4 |
| 2025-04 | 1 742 | 99.4 | 2025-01 | 40 705 | 2.0 |
| 2025-05 | 1 770 | 99.3 | 2025-02 | 40 619 | 1.9 |
| 2025-06 | 1 736 | 99.4 | 2025-03 | 38 875 | 1.9 |
| 2025-07 | 1 801 | 99.4 | 2025-04 | 38 452 | 2.0 |
| 2025-08 | 1 797 | 99.4 | 2025-05 | 38 287 | 2.3 |
| 2025-09 | 1 842 | 99.6 | 2025-06 | 39 893 | 2.4 |
| 2025-10 | 1 878 | 99.5 | 2025-07 | 40 654 | 3.8 |
| 2025-11 | 1 879 | 99.4 | 2025-08 | 41 263 | 6.1 |
| 2025-12 | 1 921 | 99.5 | 2025-09 | 42 581 | 45.2 |
|  |  |  | 2025-10 | 43 145 | 75.3 |
|  |  |  | 2025-11 | 41 453 | 95.5 |
|  |  |  | 2025-12 | 38 681 | 88.7 |

Months listed by the national quality checks for the two states in which the placeholder identifier carried a material share of records. End of data: Goiás, October 2024; São Paulo, July 2025 (5% threshold) or November 2024 (1% threshold).

**Table S16:**
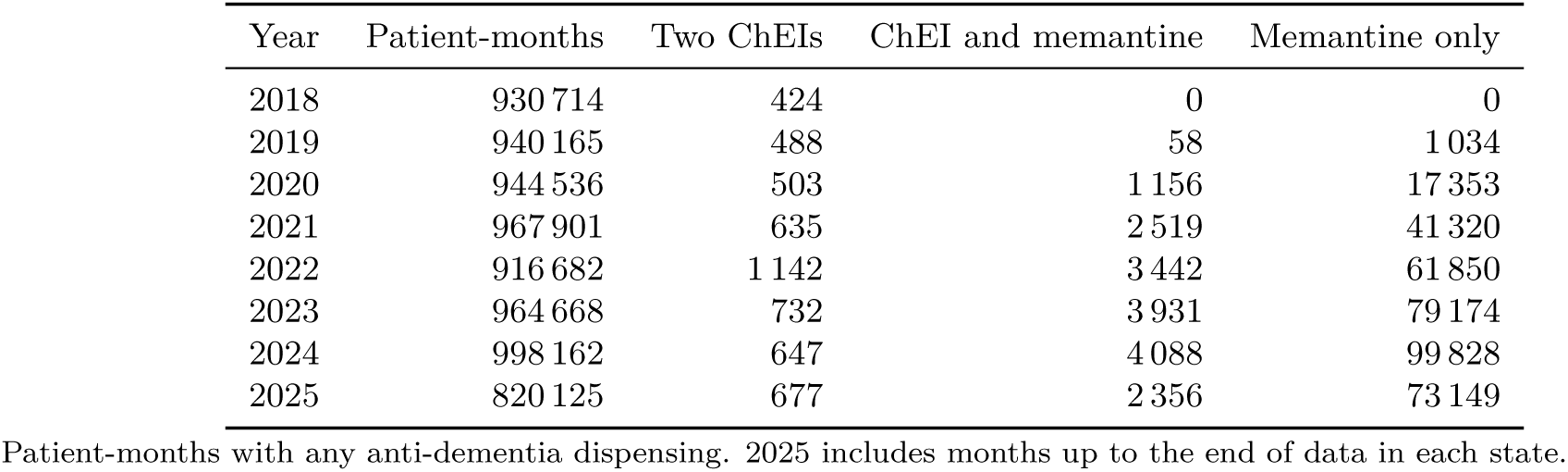
Drug combinations in the same patient-month, by year.

**Table S17:** New users by state of residence and drug, and the smallest share of any drug in each state.

| State | Donepezil | Galantamine | Rivastigmine | Total | Smallest share (%) |
|---|---|---|---|---|---|
| Acre | 9 | 21 | 27 | 57 | 15.8 |
| Alagoas | 465 | 253 | 533 | 1 251 | 20.2 |
| Amazonas | 490 | 175 | 152 | 817 | 18.6 |
| Bahia | 1 743 | 809 | 870 | 3 422 | 23.6 |
| Ceará | 2 345 | 747 | 1 184 | 4 276 | 17.5 |
| Distrito Federal | 131 | 115 | 185 | 431 | 26.7 |
| Espírito Santo | 2 010 | 1 282 | 1 418 | 4 710 | 27.2 |
| Goiás | 843 | 473 | 1 040 | 2 356 | 20.1 |
| Maranhão | 332 | 224 | 321 | 877 | 25.5 |
| Minas Gerais | 3 683 | 1 868 | 4 020 | 9 571 | 19.5 |
| Mato Grosso do Sul | 705 | 435 | 389 | 1 529 | 25.4 |
| Mato Grosso | 376 | 256 | 372 | 1 004 | 25.5 |
| Pará and Amapá | 253 | 83 | 118 | 454 | 18.3 |
| Paraíba | 628 | 740 | 863 | 2 231 | 28.1 |
| Pernambuco | 466 | 413 | 757 | 1 636 | 25.2 |
| Piauí | 515 | 239 | 285 | 1 039 | 23.0 |
| Paraná | 12 493 | 3 919 | 5 080 | 21 492 | 18.2 |
| Rio de Janeiro | 621 | 397 | 789 | 1 807 | 22.0 |
| Rio Grande do Norte | 733 | 546 | 399 | 1 678 | 23.8 |
| Rondônia | 147 | 91 | 152 | 390 | 23.3 |
| Roraima | 56 | 26 | 16 | 98 | 16.3 |
| Rio Grande do Sul | 2 866 | 1 583 | 1 392 | 5 841 | 23.8 |
| Santa Catarina | 2 905 | 1 702 | 1 472 | 6 079 | 24.2 |
| Sergipe | 160 | 88 | 199 | 447 | 19.7 |
| São Paulo | 39 150 | 14 172 | 15 657 | 68 979 | 20.5 |
| Tocantins | 40 | 34 | 212 | 286 | 11.9 |

Amapá was pooled with Pará because of the small number of new users per drug.

**Table S18:** Distribution of stabilized inverse probability of treatment weights and effective sample size, by drug.

| Drug | Mean | P1 | Median | P99 | Maximum | Maximum after truncation | Effective *n* |
|---|---|---|---|---|---|---|---|
| Donepezil | 1.00 | 0.74 | 0.94 | 1.84 | 6.12 | 1.88 | 71 230 |
| Galantamine | 1.00 | 0.56 | 0.98 | 1.67 | 2.83 | 1.88 | 29 122 |
| Rivastigmine | 1.00 | 0.44 | 0.97 | 2.07 | 4.67 | 1.88 | 34 294 |

P1, P99: 1st and 99th percentiles. Truncation at the 1st and 99th percentiles was applied to the product of treatment and censoring weights; the effective sample size refers to the truncated combined weights.

**Table S19:**
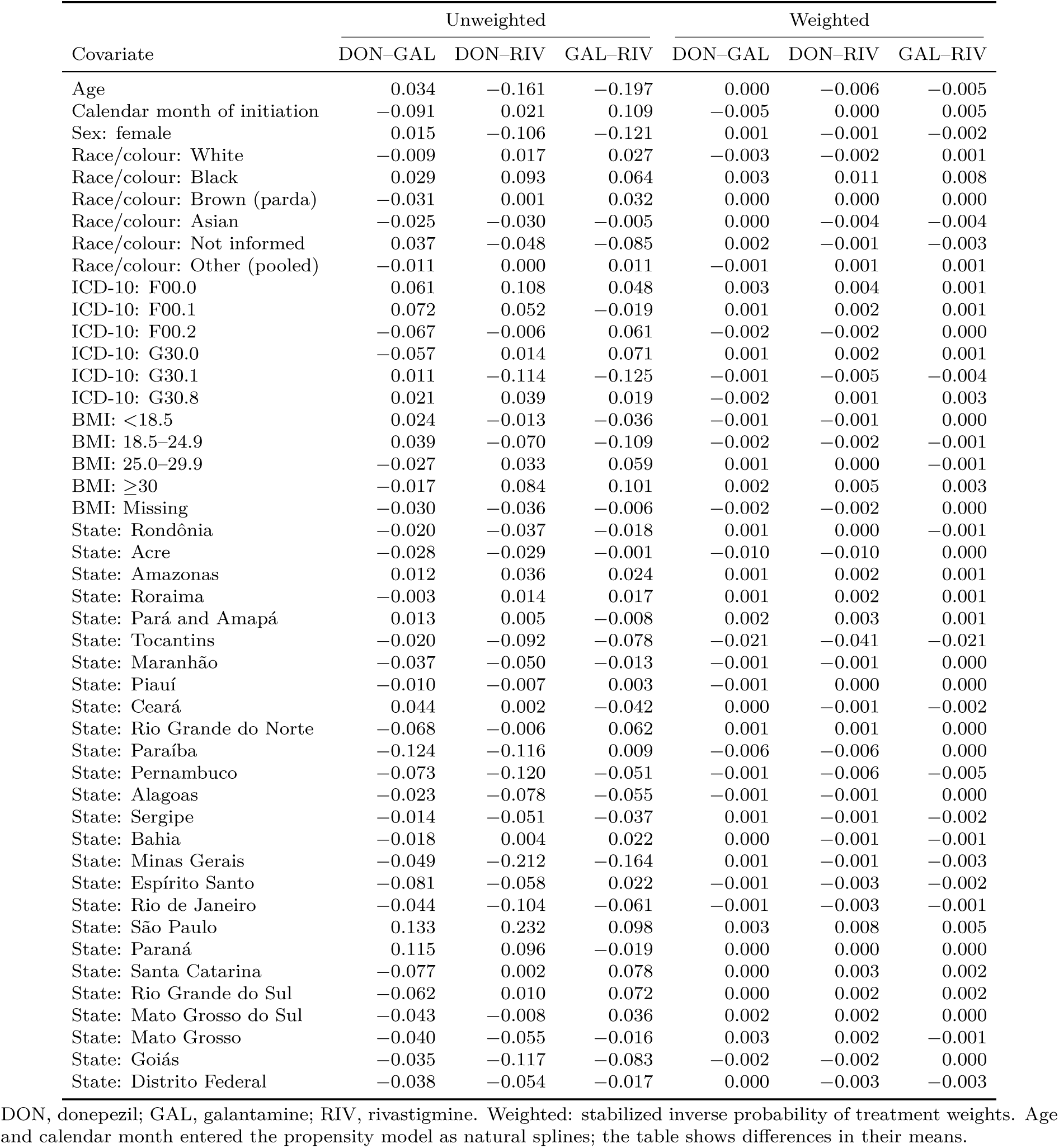
Standardized mean differences for all baseline covariates, before and after weighting, for each pairwise comparison.

|  | Unweighted |  |  | Weighted |  |  |
|---|---|---|---|---|---|---|
| Covariate | DON–GAL | DON–RIV | GAL–RIV | DON–GAL | DON–RIV | GAL–RIV |
| Age | 0.034 | −0.161 | −0.197 | 0.000 | −0.006 | −0.005 |
| Calendar month of initiation | −0.091 | 0.021 | 0.109 | −0.005 | 0.000 | 0.005 |
| Sex: female | 0.015 | −0.106 | −0.121 | 0.001 | −0.001 | −0.002 |
| Race/colour: White | −0.009 | 0.017 | 0.027 | −0.003 | −0.002 | 0.001 |
| Race/colour: Black | 0.029 | 0.093 | 0.064 | 0.003 | 0.011 | 0.008 |
| Race/colour: Brown (parda) | −0.031 | 0.001 | 0.032 | 0.000 | 0.000 | 0.000 |
| Race/colour: Asian | −0.025 | −0.030 | −0.005 | 0.000 | −0.004 | −0.004 |
| Race/colour: Not informed | 0.037 | −0.048 | −0.085 | 0.002 | −0.001 | −0.003 |
| Race/colour: Other (pooled) | −0.011 | 0.000 | 0.011 | −0.001 | 0.001 | 0.001 |
| ICD-10: F00.0 | 0.061 | 0.108 | 0.048 | 0.003 | 0.004 | 0.001 |
| ICD-10: F00.1 | 0.072 | 0.052 | −0.019 | 0.001 | 0.002 | 0.001 |
| ICD-10: F00.2 | −0.067 | −0.006 | 0.061 | −0.002 | −0.002 | 0.000 |
| ICD-10: G30.0 | −0.057 | 0.014 | 0.071 | 0.001 | 0.002 | 0.001 |
| ICD-10: G30.1 | 0.011 | −0.114 | −0.125 | −0.001 | −0.005 | −0.004 |
| ICD-10: G30.8 | 0.021 | 0.039 | 0.019 | −0.002 | 0.001 | 0.003 |
| BMI: <18.5 | 0.024 | −0.013 | −0.036 | −0.001 | −0.001 | 0.000 |
| BMI: 18.5–24.9 | 0.039 | −0.070 | −0.109 | −0.002 | −0.002 | −0.001 |
| BMI: 25.0–29.9 | −0.027 | 0.033 | 0.059 | 0.001 | 0.000 | −0.001 |
| BMI: ≥30 | −0.017 | 0.084 | 0.101 | 0.002 | 0.005 | 0.003 |
| BMI: Missing | −0.030 | −0.036 | −0.006 | −0.002 | −0.002 | 0.000 |
| State: Rondônia | −0.020 | −0.037 | −0.018 | 0.001 | 0.000 | −0.001 |
| State: Acre | −0.028 | −0.029 | −0.001 | −0.010 | −0.010 | 0.000 |
| State: Amazonas | 0.012 | 0.036 | 0.024 | 0.001 | 0.002 | 0.001 |
| State: Roraima | −0.003 | 0.014 | 0.017 | 0.001 | 0.002 | 0.001 |
| State: Pará and Amapá | 0.013 | 0.005 | −0.008 | 0.002 | 0.003 | 0.001 |
| State: Tocantins | −0.020 | −0.092 | −0.078 | −0.021 | −0.041 | −0.021 |
| State: Maranhão | −0.037 | −0.050 | −0.013 | −0.001 | −0.001 | 0.000 |
| State: Piauí | −0.010 | −0.007 | 0.003 | −0.001 | 0.000 | 0.000 |
| State: Ceará | 0.044 | 0.002 | −0.042 | 0.000 | −0.001 | −0.002 |
| State: Rio Grande do Norte | −0.068 | −0.006 | 0.062 | 0.001 | 0.001 | 0.000 |
| State: Paraíba | −0.124 | −0.116 | 0.009 | −0.006 | −0.006 | 0.000 |
| State: Pernambuco | −0.073 | −0.120 | −0.051 | −0.001 | −0.006 | −0.005 |
| State: Alagoas | −0.023 | −0.078 | −0.055 | −0.001 | −0.001 | 0.000 |
| State: Sergipe | −0.014 | −0.051 | −0.037 | 0.001 | −0.001 | −0.002 |
| State: Bahia | −0.018 | 0.004 | 0.022 | 0.000 | −0.001 | −0.001 |
| State: Minas Gerais | −0.049 | −0.212 | −0.164 | 0.001 | −0.001 | −0.003 |
| State: Espírito Santo | −0.081 | −0.058 | 0.022 | −0.001 | −0.003 | −0.002 |
| State: Rio de Janeiro | −0.044 | −0.104 | −0.061 | −0.001 | −0.003 | −0.001 |
| State: São Paulo | 0.133 | 0.232 | 0.098 | 0.003 | 0.008 | 0.005 |
| State: Paraná | 0.115 | 0.096 | −0.019 | 0.000 | 0.000 | 0.000 |
| State: Santa Catarina | −0.077 | 0.002 | 0.078 | 0.000 | 0.003 | 0.002 |
| State: Rio Grande do Sul | −0.062 | 0.010 | 0.072 | 0.000 | 0.002 | 0.002 |
| State: Mato Grosso do Sul | −0.043 | −0.008 | 0.036 | 0.002 | 0.002 | 0.000 |
| State: Mato Grosso | −0.040 | −0.055 | −0.016 | 0.003 | 0.002 | −0.001 |
| State: Goiás | −0.035 | −0.117 | −0.083 | −0.002 | −0.002 | 0.000 |
| State: Distrito Federal | −0.038 | −0.054 | −0.017 | 0.000 | −0.003 | −0.003 |

DON, donepezil; GAL, galantamine; RIV, rivastigmine. Weighted: stabilized inverse probability of treatment weights. Age and calendar month entered the propensity model as natural splines; the table shows differences in their means.

## Additional analyses

**Table S20:** Additional analyses on disease severity and unrecorded deaths: risk differences (per 100 patients) and risk ratios relative to donepezil at 24 and 36 months.

|  |  | GAL vs DON |  | RIV vs DON |  |
|---|---|---|---|---|---|
| Analysis | Months | RD | RR | RD | RR |
| *Main analysis (Table 3)* |  |  |  |  |  |
| All new users | 24 | −0.7 (−1.1, −0.3) | 0.93 (0.88, 0.97) | −4.1 (−4.5, −3.7) | 0.58 (0.55, 0.61) |
|  | 36 | −1.0 (−1.7, −0.2) | 0.93 (0.88, 0.99) | −5.5 (−6.0, −4.8) | 0.61 (0.57, 0.65) |
| *A. Composite outcome: memantine add-on or end of dispensing* |  |  |  |  |  |
| All new users | 24 | −2.2 (−2.9, −1.4) | 0.96 (0.94, 0.97) | −4.1 (−4.8, −3.3) | 0.92 (0.90, 0.93) |
|  | 36 | −2.3 (−3.1, −1.5) | 0.96 (0.95, 0.98) | −5.1 (−5.9, −4.3) | 0.92 (0.91, 0.93) |
| *B. Rivastigmine by formulation* |  |  |  |  |  |
| Oral rivastigmine | 24 | −0.6 | 0.94 | −2.7 | 0.71 |
|  | 36 | −0.9 | 0.93 | −3.0 | 0.78 |
| Transdermal rivastigmine | 24 | −0.7 | 0.93 | −4.5 | 0.54 |
|  | 36 | −0.9 | 0.93 | −6.3 | 0.55 |
| *C. Age group* |  |  |  |  |  |
| <75 years | 24 | −1.0 | 0.89 | −3.3 | 0.64 |
|  | 36 | −1.2 | 0.91 | −3.7 | 0.72 |
| 75–84 years | 24 | −0.9 | 0.91 | −4.3 | 0.57 |
|  | 36 | −0.7 | 0.95 | −6.2 | 0.56 |
| ≥85 years | 24 | 0.6 | 1.06 | −4.8 | 0.53 |
|  | 36 | −0.9 | 0.94 | −6.6 | 0.54 |

DON, donepezil; GAL, galantamine; RIV, rivastigmine; RD, risk difference; RR, risk ratio. Main analysis and A: 95% CI from 200 bootstrap replicates (percentile method). A: memantine add-on, loss to follow-up or recorded death, whichever came first, weighted by stabilized inverse probability of treatment weights (the only remaining censoring is the administrative end of data). B and C: memantine add-on with inverse probability of treatment and censoring weights, propensity model refitted in each subset; point estimates. B: oral or transdermal rivastigmine users, each analysed with all donepezil and galantamine users.

**Table S21:** Patients and events in the additional analyses, by drug.

| Analysis | Donepezil | Galantamine | Rivastigmine |
|---|---|---|---|
| A. Composite outcome | 74 165 (39 354) | 30 691 (15 125) | 37 902 (18 525) |
| B. Oral rivastigmine | 74 165 (5 479) | 30 691 (2 391) | 8 110 (487) |
| B. Transdermal rivastigmine | 74 165 (5 479) | 30 691 (2 391) | 29 792 (1 455) |
| C. Age <75 years | 26 174 (1 949) | 11 109 (858) | 11 118 (660) |
| C. Age 75–84 years | 33 943 (2 550) | 14 278 (1 125) | 17 942 (909) |
| C. Age ≥85 years | 14 048 (980) | 5 304 (408) | 8 842 (373) |

Values are new users (events). Events: composite outcome in A; memantine add-on in B and C. Cells with fewer than 5 patients are shown as <5.

